# Enhancing spontaneous recovery after stroke: A randomised controlled trial

**DOI:** 10.1101/2025.11.19.25340598

**Authors:** Winston D. Byblow, Maxine J. Shanks, Benjamin Scrivener, Laura Duval, Cathy M. Stinear, Arier Lee, P. Alan Barber, Patricia Colle, April Ren, John Cirillo, Ahmet Arac, Naveed Ejaz, Michelle Chan-Cortes, Gangadhar Garipelli, Tomoko Kitago, John W. Krakauer

## Abstract

This Phase IIa biomarker-guided stroke rehabilitation trial aimed to determine if a 3-week programme of high-dose, high-intensity (HDHI) virtual exploratory movement (VEM) therapy using MindPod Dolphin could improve upper limb recovery and outcomes early after stroke. Sixty-four participants were randomised into VEM (n=31) or conventional therapy (CoT) groups (n=33) and began intervention within two weeks of stroke. Participants were given 90 minutes of therapist time per weekday for three weeks to undertake intensive upper limb therapy over and above their usual customary care. Outcomes were obtained immediately post-intervention, three and six months post-stroke. The primary endpoint was change in Action Research Arm Test (ARAT) score between baseline and three months post-stroke. Secondary outcomes were Fugl-Meyer assessment (FM-UE), hand dexterity, reaching kinematics and transcranial magnetic stimulation-derived measures across post-intervention time points up to six months. Data from all participants were analysed for intention to treat, while fifty-four met the weekly target of active therapy minutes, permitting inclusion in a per protocol analysis. The average weekly time on task increased over the intervention period, with progressively greater distances of arm movements (VEM) or task repetitions (CoT) made each week. Still, during the final week of intervention, participants analysed per protocol spent an average of only 50 (VEM) - 66% (CoT) of the extra time available on task. For the intention-to-treat analysis, there was no effect of group for the primary endpoint. For the per protocol analysis, there was no effect of group or group by time interaction for any secondary outcome measure. An expected effect of time was observed indicative of recovery from impairment (mean ΔFM at 6 months = 23 points), increased activity capacity (mean ΔARAT at 6 months = 31 points) and improved secondary neuroscientific measures. Per protocol participants from both groups were compared to an historical cohort matched for baseline age, stroke severity and impairment who only received usual customary care. Despite three-fold difference in active upper limb therapy minutes there were no differences in the three-month ARAT or ΔFM. Indeed, the recovery in FM-UE at 3 months was proportional to baseline impairment, suggesting that the gains could be largely attributable to spontaneous biological recovery. While patient-related factors limited the dose of therapy achievable in this trial, clinician and service-related factors would also need to be overcome in routine clinical practice, to deliver equivalent doses at this early phase. It may be necessary to forego HDHI therapy until the late sub-acute phase.

## Introduction

Stroke is a leading cause of adult disability, affecting one in four adults over the age of 25.^1^ One of the main drivers of disability after stroke is motor impairment.^2^ Endogenous repair processes are considered to be largely responsible for the recovery from motor impairment that occurs early after stroke, often referred to as spontaneous biological recovery (SBR).^3–5^ However, treatments that meaningfully interact with SBR are lacking. Initial impairment and a functionally intact corticospinal tract are strongly associated with impairment recovery and outcome, independent of the time spent in upper limb therapy (i.e., therapy dose) delivered.^6–12^ One possible explanation for these findings is that therapy dose and therapy intensity are not high enough to produce the complete or near-complete motor recovery observed after experimental stroke in animals that typically make hundreds of purposeful, task-oriented movements per therapy session.^13–15^ Another possibility is that conventional stroke rehabilitation may over-emphasise compensatory approaches, which may not interact with endogenous processes responsible for SBR across the sub-acute phases.^16^ These findings are challenging because they indicate that stroke rehabilitation as currently delivered may not interact effectively with processes responsible for early motor recovery.^3,7,10–12,17,18^ Novel technologies that more readily permit high-dose, high-intensity (HDHI) therapy to be offered among the many competing priorities of early sub-acute stroke rehabilitation are required.

Experimental studies of stroke and motor recovery in animals have identified a sensitive period in the initial days after injury when the neural circuitry is especially responsive to training.^19,20^ Human patients also exhibit a sensitive period of accelerated recovery after stroke.^21^ In the CPASS trial, patients had a clinically significant increase in upper limb activity compared to controls receiving standard therapy when twenty hours of additional therapy were initiated in the late sub-acute phase (30-90 days poststroke), but this was not seen when additional therapy was initiated in the chronic phase (>180 days post-stroke).^22^ Fine-grained assessment of 2D-planar reaching kinematics across the sub-acute phase also indicates that recovery plateaus within 1-2 months post-stroke, indicating a narrow timeframe for potentially interacting with SBR.^23^ Delivery of HDHI therapy at the early sub-acute phase (7-30 days^24^) is expected to have the best possible chance of interacting with endogenous processes responsible for SBR.^25^

An emerging view in stroke rehabilitation research is that new interventions should contrast highly with conventional therapy to detect a clinically meaningful effect.^4^ The SMARTS2 study examined the efficacy of a novel video-game-based therapy on upper limb recovery at the sub-acute phase after stroke . The game is driven by a physics-based engine that creates new challenges in a fun and rewarding environment to tease out large amplitude and high-quality non-synergistic movement patterns allowing exploration of the full workspace personalised to the player (patient) based on their current active range of movement in 3D. The SMARTS2 trial found no difference between the intervention and time-matched conventional therapy on upper limb impairment or activity capacity even though compliance with active therapy was high for both groups. Still, both groups improved activity capacity relative to an historical control group that received usual customary care (not high dose). In SMARTS2, participants began the intervention on average 6 weeks post-stroke, when some patients have already reached their motor recovery plateau. Another smaller feasibility study with MindPod Dolphin (MindMaze SA, Lausanne) was conducted earlier at the sub-acute phase, consisting of up to 40 sessions, on top of usual customary care in a rehabilitation centre.^26^ Therapy was started on average 32 days post-stroke, and deemed feasible with patients completing on average up to 33 of 60 minutes of treatment using MindPod Dolphin . These findings demonstrate the feasibility of conducting a high-dose, high-intensity randomised controlled trial early after stroke.

The “Enhancing Spontaneous Recovery after Stroke Study” (ESPRESSo) is a single-site, randomised, assessor-blind, controlled Phase IIa clinical trial. ESPRESSo was designed to evaluate whether HDHI virtual exploratory movement therapy (VEM), an impairment-oriented rehabilitation approach that promotes self-generated exploratory upper limb movements within a virtual environment, starting within 2 weeks of stroke, can enhance spontaneous recovery over and above time-matched additional conventional upper limb therapy. Although ESPRESSo shares similarities with SMARTS2, it has two essential differences: the earlier intervention period and biomarker guidance for patient selection. While a randomised controlled trail (RCT) may overcome rehabilitation service-related barriers to delivery,^27^ patient-factors such as fatigue, motivation and cognitive efficiency may limit the extent of HDHI delivery, and these were monitored and assessed throughout the intervention phase of the trial. Biomarker guidance is critical since upper limb impairment recovery relies on a functionally intact corticospinal tract (CST). Recent consensus statements for stroke rehabilitation espouse the importance of patient selection for trials informed by biomarkers that accurately differentiate between recovery phenotypes. In particular, motor evoked potential (MEP) status is a biomarker of residual CST integrity that can be obtained early after stroke and used to minimise heterogeneity in studies of upper limb rehabilitation trials.^4,25^ As such, ESPRESSo only recruited patients with positive upper limb motor-evoked potential (MEP+) status, indicative of a functionally intact CST. While MEP+ is not necessarily a guarantee of a good functional outcome, excluding patients without MEPs reduces between-group heterogeneity.^28^ This biomarker-guided approach also improves trial efficiency by permitting inclusion of patients who have latent capacity for good upper limb recovery but are often excluded from participation owing to their baseline impairment.

The primary hypothesis was that recovery of upper limb motor capacity, measured as change in Action Research Arm Test (ARAT) score from baseline to three months post-stroke, would be better for the group that underwent early intense VEM than the group that received additional therapist-time matched conventional therapy (CoT). Secondary hypotheses compared groups in their six month recovery and outcome, recovery from upper limb motor impairment measured on the Fugl-Meyer Upper Extremity (FM-UE) assessment, recovery of upper limb motor control in terms of reaching kinematics, recovery of manual dexterity, and recovery of corticospinal excitability obtained from transcranial magnetic stimulation at one, three and six months post-stroke.

To explore the effect of early HDHI therapy irrespective of the way in which it was delivered, gaming versus extra conventional therapy, we compared upper limb recovery and outcomes of both HDHI groups to an historical cohort who received standard and usual care within the same rehabilitation setting. By design, we expected that the active time spent engaged in upper limb therapy would be greater for patients in ESPRESSo than those who received usual customary care (UCC). Upper limb recovery and outcome of ESPRESSo participants were compared to those who received UCC to better understand the interaction of early intense therapy with endogenous repair processes. This historical cohort’s baseline, recovery and outcome measures were acquired from earlier prospective observational studies.^29,30^

## Materials and methods

### Participants

Consecutive patients were screened upon admission to the Stroke Unit at Auckland City Hospital between 1 February 2021 and 31 March 2024 as part of registered clinical trial (ACTRN12620000871943). Inclusion criteria were: ≥18 years old; upper limb weakness; FM-UE score of <51; able to sit out of bed to participate in upper limb therapy; a functionally intact corticospinal tract (CST) determined by Shoulder Abduction-Finger Extension (SAFE) score of >4 within 3 days, or the presence of motor evoked potentials (MEPs) in the paretic extensor carpi radialis or first dorsal interosseous muscle within 1 week of stroke; and able to begin therapy within two weeks of ischaemic or haemorrhagic stroke confirmed by a neurologist.^31^ Patients were considered MEP+ if at least two MEPs of any amplitude were observed while the participant was at rest or maximally activating both upper limbs, with stimulation up to 100% maximum stimulator output (MSO, See Supplementary Material). Exclusion criteria were: inability to give informed consent due to cognitive or communication difficulties; cerebellar stroke; previous physical or neurological impairments that would interfere with therapy or assessments of upper limb; residing out of area; medical, social and/or personal circumstances that would preclude completing therapy or assessments; life expectancy <12 months. All participants gave written informed consent in accordance with the declaration of Helsinki. Aphasia-appropriate study information was available to support participation.

### Randomisation

Participants were randomised 1:1 to either VEM or CoT using custom software (www.rando.la). While no characteristic was prioritised to achieve balance, group differences were minimised for age, baseline stroke severity measured with the National Institutes of Health Stroke Scale (NIHSS), baseline FM-UE, concordance and whether reperfusion therapy was received.

### Interventions

All participants received usual customary care (physiotherapy and occupational therapy) from inpatient and community rehabilitation teams throughout the trial. Participants were either seated or standing, depending on their ability, fatigue and the requirement for engaging in the task. Intervention sessions were integrated into the participant’s standard multidisciplinary therapy schedule. (See Supplementary Material). The interventions for both groups were designed to achieve high-dose, by maximising the total time of upper limb therapy, and high-intensity, by maximising movement repetitions completed per session, as much as practicable at the early sub-acute phase. Briefly, participants were allotted 90 minutes of therapist time for each intervention session. For adherence to the therapy protocol, participants were required to complete a minimum of 360 minutes of therapist time in their first week five sessions (week 1), 390 minutes in the next five (week 2), and 420 minutes in the final five (week 3). Therapists reinforced the expectation to perform as much active upper limb therapy (i.e., time on task) as possible during each session and used a stopwatch to record active upper limb therapy to the minute. Patients reported their motivation at the start of each session (strongly, motivated, slightly, or not motivated), and their fatigue on a 10-point scale (0 = nil, 10 = worst) at the start and end of each session. Also, in each intervention session, a rapid assessment of cognitive efficiency was obtained using the DANA Brain Vital during a rest break (See Supplementary Material).

### Virtual Exploratory Movement Therapy (VEM)

The VEM intervention involved an immersive video game-based, animated, neurotherapeutic platform called MindPod Dolphin (MindMaze SA, Lausanne).^32^ The MindPod platform captures the participant’s paretic arm movement through a markerless motion capture system (Microsoft Kinect V2 sensor). It provides visual feedback such that arm movement made by the participant is reflected through the swimming of a virtual dolphin or whale. Tasks and gameplay are designed to promote movement in all planes and are titrated based on the successful completion of progressive difficulty levels. During these “arm” levels, the MindPod platform calculates time spent actively moving the paretic limb in minutes and the metric distance travelled by the paretic hand. This information is displayed to participants to motivate their progression. For hand-specific activities, a custom an air-sealed hand-held instrumented controller called Izar (MindMaze SA, NeuroX Group SA, Lausanne) housed with sub-newton pressure and inertial sensor, was used to control gameplay. The “hand” levels of MindPod involve a variety of tasks controlling aquatic creatures through force grading, timing, and endurance, as well as pinch, grasp, and multi-joint coordination with the fingers and wrist. Therapists provided verbal and tactile feedback to reinforce high-quality movements, game progression and exploration of the full workspace. (Further details in Supplementary Material).

### Conventional Therapy (CoT)

The additional time-matched conventional therapy (CoT) was administered in alignment with the principles of upper-extremity task-specific training.^33^ This included individualised goal setting during the initial session and a selection of therapy tasks that aligned with these goals. Tasks typically simulated functional activities of daily living such as dressing, cooking, eating, grooming and cleaning. An inertial movement sensor (IMU, Vicon Motion Systems, Oxford United Kingdom) was placed on the paretic wrist to measure the time spent actively moving the upper limb. All tasks were documented by the intervention therapist, including the amount of assistance provided, total repetitions, time spent actively on task, rest time, any modifications, and total time spent on each task (Further details in Supplementary Material).

### Blinding and Assessments

Trained clinical assessors acquired all post-intervention assessments blinded to group allocation. Regular training sessions were held to ensure consistency between blinded assessors (See Supplementary Material). At baseline, clinical assessments were obtained within the first two weeks after stroke and within one day of the start of the intervention whenever possible. Baseline assessments included the NIHSS, Charlson Comorbidity Index (CCI), FM-UE, and ARAT. The FM-UE assessment measures motor impairment, with the upper extremity portion focusing on movements in and out of synergy and compensatory movements. The maximum score is 66, with higher scores reflecting lower impairment.^34^ The ARAT assesses upper limb activity capacity through grasp, grip, pinch and gross movements with a total score of 57 and higher scores reflecting better function.^35^ The FM-UE and ARAT, as well as neurophysiological, kinematic and manual dexterity assessments, were then obtained at three post-intervention timepoints: immediately post-intervention (4 – 5 weeks post-stroke, herein referred to as one-month assessment), three months and six months post-stroke. The modified Rankin Scale (mRS) and Stroke Impact Scale version 3.0 (SIS) were obtained at six-month follow-up only. The mRS evaluates global disability,^36^ and the SIS is a stroke-specific, patient-reported measure assessing multidimensional stroke outcomes and activity limitations.^37^ An SIS score of 100% indicates that the stroke has had no lasting impact on the patient’s life. During the rest break of each intervention session, reaction time performance was assessed on tasks presented via a smartphone application (DANA Brain Vital V3.0.8) to generate a cognitive efficiency score for the participant during each therapy session. A subset of participants who received VEM rated their enjoyment.^38^ (Further details in Supplementary Material).

Additional neuroscientific assessments were obtained at all post-intervention timepoints by non-blinded experimenters for dexterity, reaching and neurophysiology as described below. Assessments were made from the non-paretic side first, then the paretic side.

### Manual Dexterity

Manual dexterity was assessed during a visuomotor force tracking task of a sinusoidal waveform, while holding an Izar device. The hand-held Izar device is roughly egg-shaped, measuring approximately 61 x 82 mm with a flat base and silicone rubber gripping surface. The target and produced force were displayed to the participant using custom-built software (MindMaze SA, Lausanne). Visuomotor tracking was performed by grasping the device with the whole hand, or pinching using a precision grip between the thumb and index finger. Maximum voluntary force (MVF) was calculated from the largest of three maximal squeeze attempts with each type of grip. Nine trials were collected from each hand with grasp and then pinch, consisting of three trials at 20%, 40%, and 60% of MVF, in a randomised order. During each trial, three squeeze and release actions were required to accurately track a sinusoidal force profile displayed on a screen. Each squeeze and release involved a 5 s ramp-up to the target force and 5 s ramp-down to zero force. Rest periods were offered between trials to prevent fatigue.

### Reaching Kinematics

Reaching was performed with the participant seated at a table with a clear acrylic top. The table was fitted with five evenly spaced rods, the tops of which were reaching targets at predetermined coordinates.^39^ Participants started each reach with the shoulder in a neutral position, the arm resting alongside the body, the elbow flexed at 90°, with their hand placed on the table in the midline. Reaches were made to each of the five targets in sequential order. The first target was directly in front of the midline. Targets two and three were positioned on the same side as the reaching hand. Targets four and five require reaching across the midline.

Despite the different directions and heights of the targets, the reach distance was a uniform length from the starting position for all five targets. Participants were instructed to prioritise smoothness over speed. The five-target sequence was repeated ten times for each upper limb. The reaches were captured using a markerless infrared video-capture system.^40^ (See Supplementary Material).

### TMS-derived Stimulus-Response Curves

TMS was applied with a MagPro X100 stimulator with Option, with a monophasic pulse directed to induce posterior-anterior current in the brain in Power mode (MagVenture, Farum, Denmark). The experimenter held a figure-of-eight coil (MC-B70) over the M1 hand area with the handle posterolateral at approximately 45 degrees from the midline. An optimal position for eliciting MEPs in surface EMG of four contralateral muscles was determined and marked on the head to ensure consistent coil placement throughout the session and continuously monitored to ensure it was held precisely at the marked hotspot. Stimulation intensities ranged from 30 – 100% MSO in 10% increments, as well as 65% MSO to increase sensitivity near the midpoint of the stimulus-response (S-R) curve. (See Supplementary Material).

### Data processing and dependent measures

#### Manual Dexterity

Data were processed using custom MATLAB scripts (MATLAB R2023a). Dexterity was evaluated by calculating the root-mean-square error (RMSE) between the participant’s generated trace and the sinusoidal trace they were tracking. The average RMSE across the three force levels was computed to measure dexterity for each hand while performing with pinch or grasp.

#### Reaching Kinematics

The recordings from the two cameras produced two synchronised videos, which were processed using OpenPose (version 1.4.0) to detect joint positions. The corresponding 2D joint positions from each video were then stereo triangulated to estimate their 3D locations. The wrist velocity profile and trunk displacement angle were extracted for each reach. The Spectral Arc Length (SPARC) of the time-normalised wrist velocity profile was calculated using a custom script (Python) to determine reaching smoothness.^41^ Trunk tilt angles were extracted for each target, averaged, and then summed across targets. This approach accounted for variations in target distance and height, preserving the distinct demands of each target instead of treating them uniformly. (See Supplementary Material).

#### TMS-derived Stimulus-Response Curves

The average MEP amplitude for each muscle was plotted as a function of TMS intensity, as in previous studies,^42,43^ by determining a best-fitting sigmoid function using MATLAB. MEPsum was calculated as the area under the curve adjusted for background EMG amplitude and used as the main measure of corticospinal excitability. Estimated resting motor threshold (eRMT) was calculated as the first stimulation intensity where the sigmoid function consistently exceeded 50 μV. Group average S-R curves were constructed for the paretic and non-paretic hand at each time point. (See Supplementary Material).

#### Lateralisation Indices

Manual dexterity, kinematic and neurophysiology measures were represented and analysed as lateralisation indices calculated as:

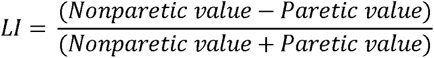

The lateralisation index (LI) reflects a normalised difference between the non-paretic and paretic sides. For MEPsum and trunk tilt values, a positive LI reflects a performance favouring the non-paretic side, while a negative LI favours the paretic side. For RMSE, eRMT, and SPARC values a positive LI reflects a performance favouring the paretic side, while a negative LI favours the non-paretic side. For all measures, an LI of 0 indicates equivalence between non-paretic and paretic sides.

#### Statistical analyses

The study was powered to detect a difference of 7 points on the ARAT, slightly larger than the minimal clinically important difference (MCID = 5.7). The estimated sample size required to detect this effect with 90% power was 106 patients. Statistical analyses were conducted using SAS version 9.4. and GraphPad Prism version 10. The primary endpoint was ΔARAT at three months. Secondary outcomes included ΔARAT at one and six months, ΔFM at one, three, and six months post-stroke, and mRS and SIS score at six months post-stroke.

Neuroscientific measures related to dexterity, reaching kinematics and corticospinal excitability were assessed at all post-intervention time points. Mixed model for repeated measures (MMRM) was used for all primary and secondary analyses involving repeated measurements. Fixed effects included Group (VEM, CoT), Time (one, three, six months), Group x Time interaction, baseline ARAT score and days post-stroke for baseline assessment. Randomisation variables of age, baseline NIHSS and FM-UE, hand concordance (Y/N), and reperfusion therapy (Y/N) were included as fixed effects. Median mRS and SIS scores were analysed using the Mann-Whitney U test. Intention-to-treat analyses considered all randomised participants regardless of whether the therapy protocol was met or follow-up assessments were obtained. Per protocol analyses were used to explore the effect of Group, Time and Group x Time with greater sensitivity to determine the effect of impairment-oriented VEM versus task-oriented CoT for participants who adhered to the HDHI therapy protocol. Unless stated otherwise, least squares mean estimates and 95% confidence intervals are reported. Two-sided p<0.05 was used to determine statistical significance.

#### Comparison to Historical Cohort

To determine the effect of early HDHI therapy on upper limb outcome and recovery, three-month ARAT and ΔFM scores were compared between the ESPRESSo *per protocol* cohort and a historical cohort (HC). The HC were selected from two observational studies carried out between 2012 and 2021 at the same hospital as the present study. The same inclusion criteria were applied while also matching age, baseline NIHSS, and baseline upper limb impairment as closely as possible. The selection was performed by a research assistant who was blinded to the three-month ARAT and FM-UE scores, with participant matching criteria confirmed independently by two experimenters (BS and WB). The HC received usual and customary stroke rehabilitation care (UCC) from allied health team members within the hospital service. Therapy time was documented in minutes per day as dictated by the observational trials. *Per protocol* participants from both groups (VEM and CoT) were pooled to create a sample who received early HDHI upper limb therapy (HDHI). An independent samples t-test was performed to compare the amount of upper limb active therapy time in minutes between the HDHI and HC groups. Multiple linear regression models were used to compare the cohorts for three-month ARAT outcome and impairment recovery (ΔFM_3M_), adjusting for baseline variables of age, NIHSS, FM-UE, days post-stroke, concordance and reperfusion therapy. Pearson correlation explored association between ΔFM_3M_ and Total Upper limb therapy time. ΔARAT could not be determined due to the absence of baseline ARAT scores for HC.

#### Recovery phenotype analysis

Recoverers and non-recoverers were differentiated, and non-recoverers were removed from the dataset to permit analysis of recovery phenotype with multiple linear regression. Non-recoverers were estimated with hypothesis-free *k-means* clustering to differentiate recovery and non-recovery phenotypes based on ΔFM at 3 months.^6^ Clustering was performed based on orthogonal distances with k = 2 or more clusters. This method minimises within-cluster variance while maximising the between-cluster variance. Once recovery phenotype patients were identified, ΔFM_3M_ data were analysed using multiple linear regression model adjusting for baseline variables as previous, and using linear regression as a special case of the formula R = β I + C, where R = ΔFM_3M_, I is initial impairment (66 – baseline FM-UE), and β is the coefficient of interest (slope), with the constant C forced to 0.^10^ F-tests were used to examine differences in slope between the HDHI and HC cohorts versus a single model for both cohorts (null hypothesis). Model fit was determined using Akaike Information Criterion corrected (AICc) for small samples to determine the preferred approach.

## Results

### Baseline Characteristics and Protocol Adherence

Participants were screened on admission to the stroke service at Auckland City Hospital between Feb 2021 and March 2024 (Fig. 1), and 64 participants were recruited and randomised into VEM or CoT (TablelJ1). The intention-to-treat (ITT) analyses contained data for all 64 participants. Of the participants randomised, 54 completed the therapy protocol and were assessed at follow-up and were included in the *per protocol* (PP) analysis. Adherence to the assessment schedule for the PP cohort was evident from the median (range) days for post-intervention at the one month [VEM = 33 (27 – 56); CoT = 33 (28 – 60)], three month [VEM = 85 (77 – 104); CoT = 87 (81 – 128)] and six month time point [VEM = 189 (178 – 320); CoT = 188 (178 – 281)]. Unfortunately, the timing of some assessments were delayed due to restrictions during the Covid-19 pandemic and/or participant illness.

**Figure 1.**
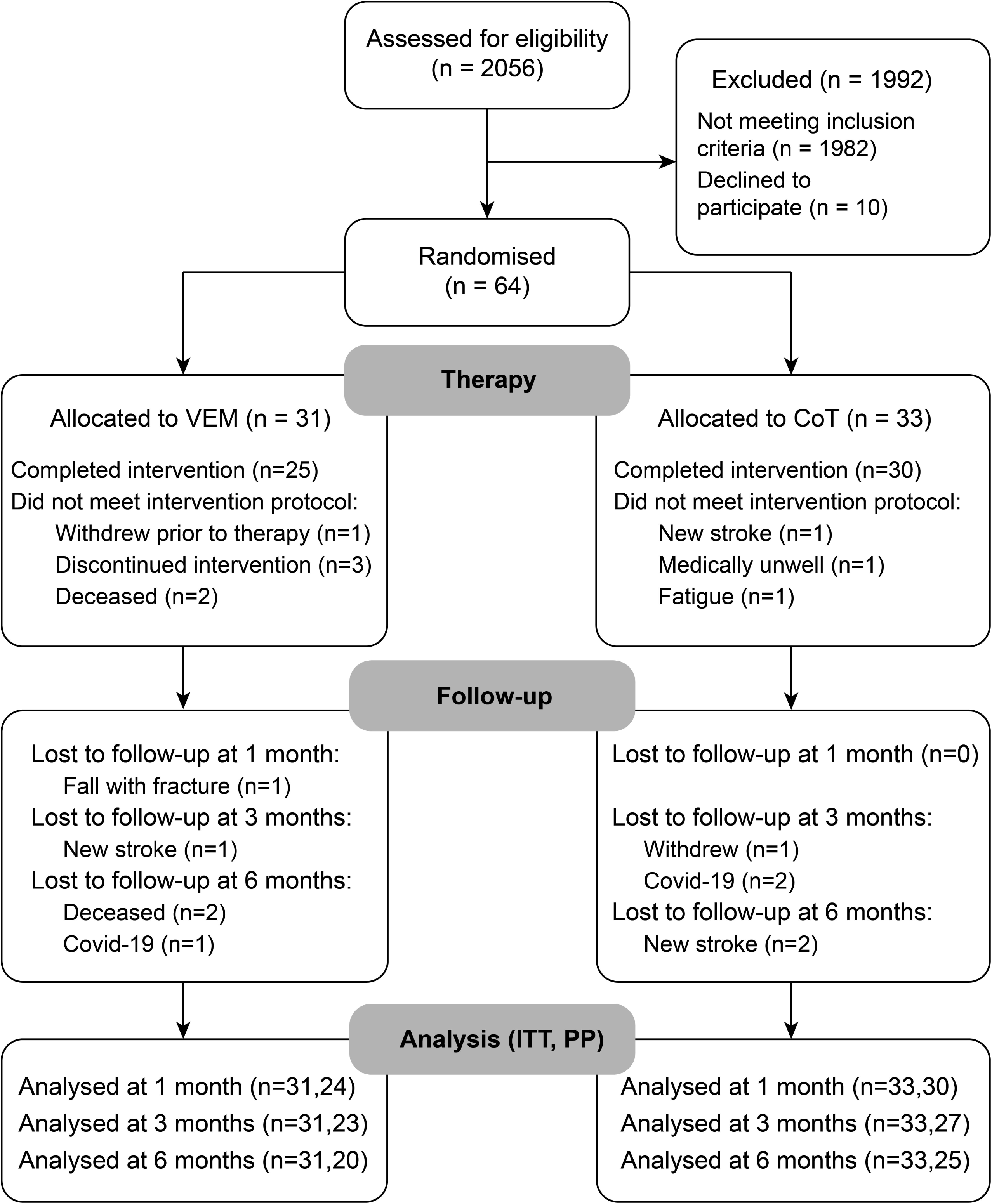
Trial Profile. VEM Virtual Exploratory Movement Therapy delivered with MindPod Dolphin; CoT Conventional Therapy; ITT Intention to Treat; PP per protocol

Intervention engagement is shown in Table 2. Several aspects were notable. First, the average weekly time on task increased over the intervention period, with progressively greater distances of arm movements (VEM) or task repetitions (CoT) achieved. Second, participants across both groups, reported high levels of motivation and increased fatigue within each of the fifteen sessions, supporting the high-intensity nature of the protocol. Cognitive efficiency also increased over the intervention period (Figure.S1). The VEM intervention platform was considered enjoyable (See Supplementary Figure 2 for further details).

Results from statistical analyses indicate per protocol data, unless otherwise indicated. Complete result tables for PP and ITT analyses can be found in the Supplementary Material.

### Primary Endpoint

The intention to treat analysis for ΔARAT scores showed a main effect of Time (*F*_2,68.2_ = 33.36, *P <* 0.0001), no effect of Group (*F*_1,46.6_ = 0.49, *P =* 0.49) and no Group x Time interaction (*F*_2,68.1_ = 0.12, *P =* 0.89). The estimated ARAT score immediately post-intervention was higher than baseline (ΔARAT = 20.2, 16.4 – 23.9, *P <* 0.0001), with a further increase between one and three months (ΔARAT = 6.1, 4.0 – 8.2, *P <* 0.0001) and between three and six months (ΔARAT = 3.0, 0.8 – 5.2, *P =* 0.01). This analysis indicates no differences between VEM and CoT at the primary endpoint, but did indicate recovery over time across both groups. Sensitivity was explored further through per protocol analyses.

### Secondary Outcomes

#### Clinical Measures

ARAT scores and least squares mean estimates of ΔARAT from per protocol analyses are shown in Fig. 2A & B respectively. There was a main effect of Time (*F*_2,66.3_ = 31.76, *P <* 0.0001), no effect of Group (*F*_1,44.4_ = 0.60, *P =* 0.44) and no Group x Time interaction (*F*_2,66.2_ = 0.18, *P =* 0.84). The estimated ARAT score immediately post-intervention was higher than baseline (ΔARAT = 20.4, 16.6 – 24.3, *P <*0.0001) with a further increase between one and three months (ΔARAT = 6.0, 3.9 – 8.1, *P <* 0.0001) and from three to six months (ΔARAT = 3.0, 0.7 – 5.2, *P <* 0.01). Analysis of baseline variables indicated significant fixed effects for baseline ARAT and FM-UE for both PP and ITT analyses, and Concordance for the ITT analysis (*P =* 0.0605 for PP analysis; see Supplementary Tables 1 and 2).

**Figure 2.**
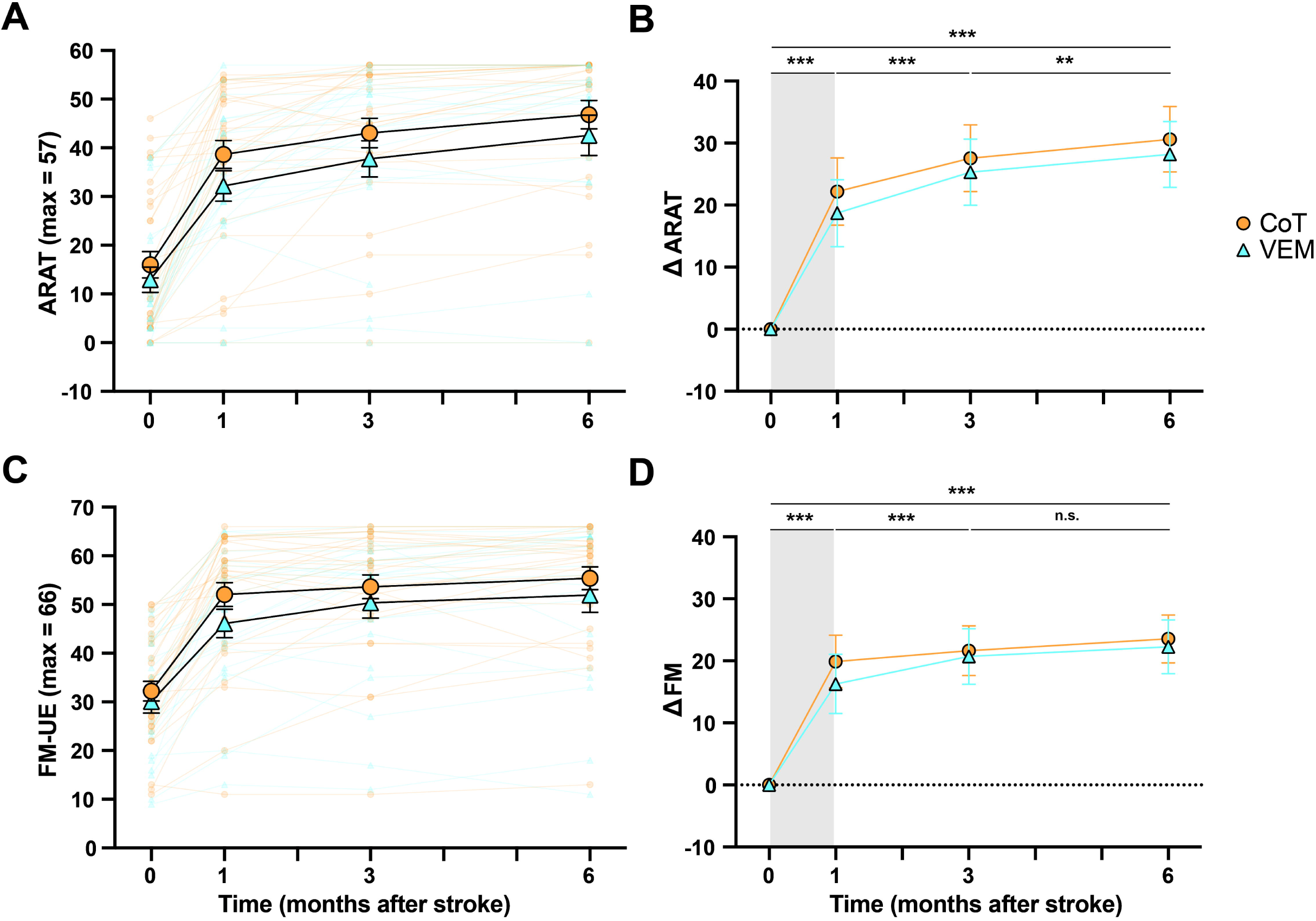
Upper-limb activity and impairment scores and model estimates from per protocol analysis. ARAT (A) and FM scores (C) for all participants (N=54) along with mean ± standard error. Least squares mean estimates and 95% confidence interval (CI) for ΔARAT (B) and ΔFM (D) from Mixed Models with Repeated Measures. Grey shading indicates the intervention period. Baseline assessment (Time 0) was obtained within 2 weeks post-stroke (see Table 3). See Supplementary Table 1 and 3 for number of samples per group and time and F-test statistics. ***p<0.001 n.s. non-significant.

FM-UE scores and least squares mean estimates of ΔFM from per protocol analyses are shown in Fig. 2C & D respectively. There was a main effect of Time (*F*_2,55.8_ = 10.2, *P <* 0.0005), no effect of Group (*F*_1,45.3_ = 0.45, *P =* 0.50) and no Group x Time interaction (*F*_2,55.8_ = 1.18, *P* = 0.31). Estimated ΔFM score immediately post-intervention was higher than baseline (ΔFM = 18.1, 14.9 – 21.2, *P <* 0.0001) with a further increase between one and three months (ΔFM = 3.1, 1.4 – 4.8, *P <* 0.001), but no further gain that was statistically significant between three and six months (ΔFM = 1.7, -0.1 – 3.5, *P =* 0.06). Analysis of baseline variables indicated a significant fixed effect for baseline FM-UE, but no other variables. The model would not converge with Concordance and Reperfusion Therapy included and these variables were removed. (See Supplementary Tables 3 and 4).

For other secondary clinical outcomes, there was no difference between groups for mRS scores (VEM = 2.5, range = 1 - 6, *n* = 22; CoT = 2, range = 1 - 4, *n* = 27; *P =* 0.30) and no difference between groups for SIS scores (VEM = 71.3%, range = 39.1 – 97.9%, n = 20; CoT = 78.9%, range = 31.4 – 94.1%, n = 26; *P =* 0.39) at six months post-stroke, with similar outcomes for the ITT analysis (both *P* > 0.35).

#### Manual Dexterity

When performing the visuomotor force tracking task using a whole hand grasp (Fig. 3A & B), there was a main effect of Time (*F*_2,78.4_ = 3.36, *P* < 0.05), no effect of Group (*F*_1,41.1_ = 0.28, *P* = 0.60) and no Group x Time interaction (*F*_2,78.4_ = 0.59, *P* = 0.56) for the LI of RMSE. LI improved between one and three months (ΔLI = -0.057, -0.109 – -0.006, *P* < 0.05) without further statistically significant improvement between three and six months post-stroke (ΔLI = -0.004, -0.058 – 0.050, *P* = 0.88). LI was initially less than zero (1m LI = - 0.101, -0.143 – -0.059, *P* < 0.0001, 3m LI = -0.044, -0.087 – -0.001, *P* < 0.05) but was not statistically different from 0 at six months post-stroke (LI = -0.040, -0.084 – 0.004, *P* = 0.07) indicating paretic side recovery to near non-paretic levels for whole hand grasp.

**Figure 3.**
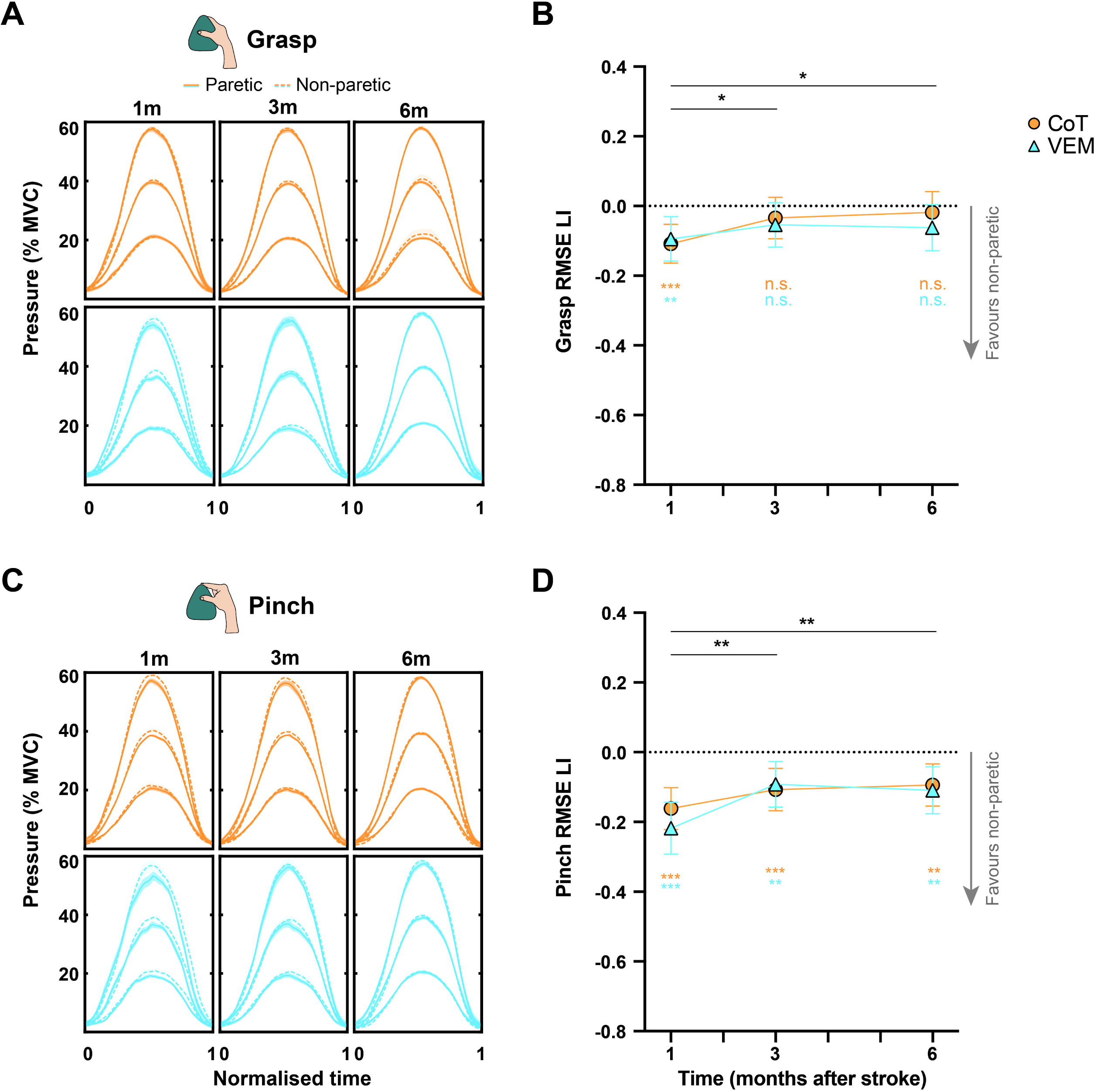
Manual dexterity performance using whole hand grasp or pinch grip and model estimates from per protocol analysis. Group average force profiles for Grasp (**A**) and Pinch (**C**). Shading denotes 1 SE. Model estimates and 95%CI from Mixed Models with Repeated Measures of RMSE LI for Grasp (**B**) and Pinch (**D**). Coloured asterisks denote significant differences from 0 from two-tailed one-sample t-tests. See Supplementary Table 5 for the number of samples per group and time and F-test statistics. *p<0.05 **p<0.01 ***p<0.001 n.s. non-significant

When performing the task using only pinch grip (Fig. 3C & D), there was a main effect of Time (*F*_2,72.8_ = 6.94, *P* < 0.005), with no effect of Group (*F*_1,37.2_ = 0.29, *P* = 0.59) or Group x Time interaction (*F*_2,72.8_ = 0.90, *P* = 0.41) for the observed RMSE expressed as LI. LI improved between one and three months (ΔLI = -0.090, -0.143 – -0.037, *P* < 0.005), with no further improvement between three and six months (ΔLI = 0.002, -0.051 – 0.055, *P* = 0.94). Pinch LI values remained below 0 across all time points (1m = -0.190, -0.237 − -0.143, *P* < 0.0001, 3m = -0.100, -0.144 – -0.056, *P* < 0.0001, 6m = -0.102, -0.146 − -0.0569, *P* < 0.0001) indicating pinch grip with paretic side remained worse than the non-paretic side.

Fixed effects of baseline variables for both analyses are shown in Supplementary Table 5.

#### Reaching Kinematics

Analyses of reaching smoothness (Fig. 4A & B) indicated a main effect of Time (*F*_2,64.5_ = 6.09, *P* < 0.005), no effect of Group (*F*_1,41.3_ = 1.02, *P* = 0.32) and no Group x Time interaction (*F*_2,64.5_ = 0.14, *P* = 0.87). Smoothness LI increased between one and three months (ΔLI = 0.025, 0.010 – 0.040, *P* < 0.005), with no further improvement from three to six months post-stroke (ΔLI = -0.002, -0.0131 – 0.0169, *P* = 0.80). At all timepoints, LI was less than 0 (1m LI = -0.061, -0.078 − -0.044, *P* < 0.0001, 3m LI = -0.036, -0.052 – -0.019, *P* < 0.0001, 6m LI = -0.038, -0.054 − -0.0214, *P* < 0.0001) indicating that reaches were made more smoothly with the non-paretic than paretic side, and early improvements on paretic side plateaued after three months.

**Figure 4.**
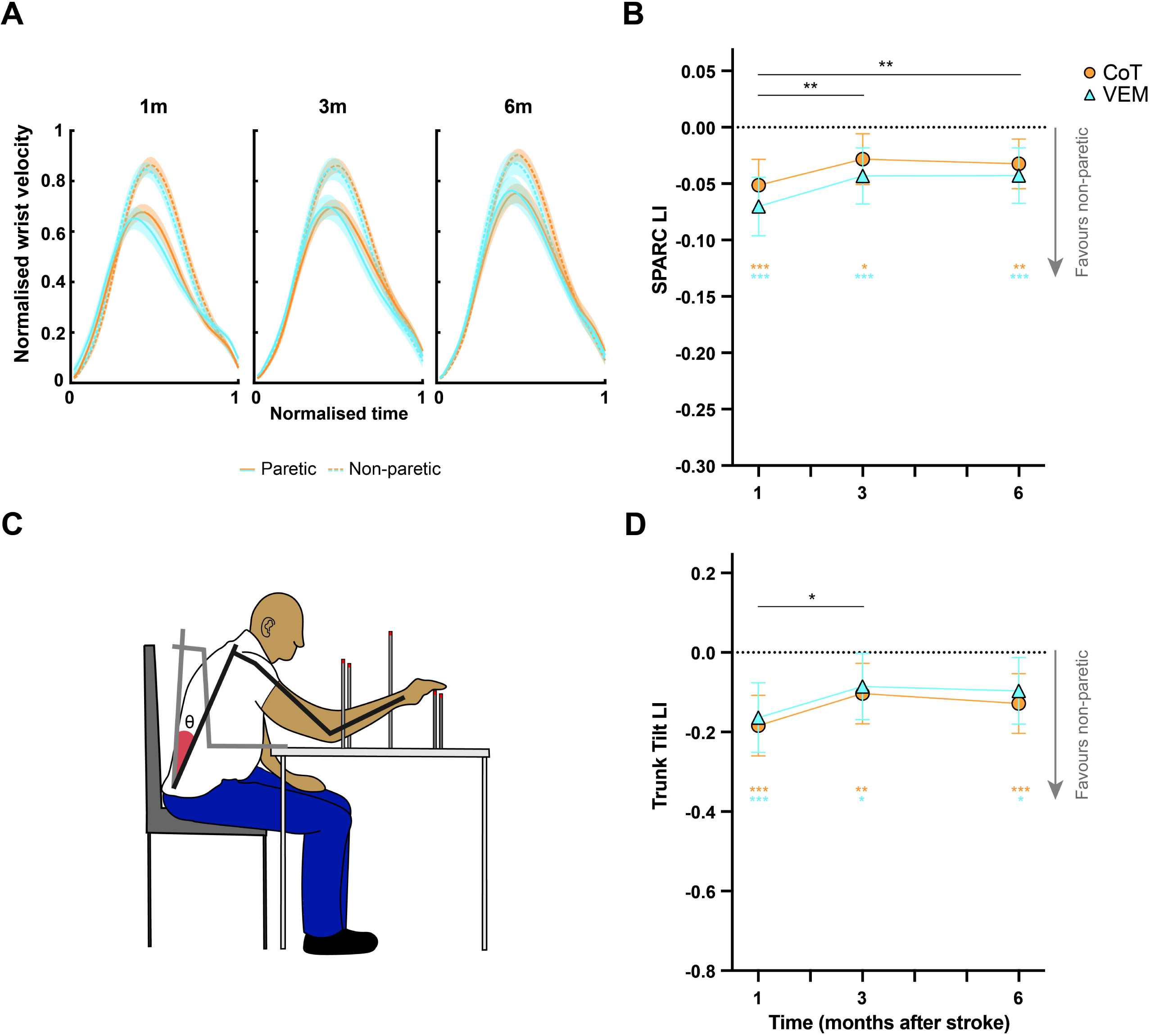
Kinematic analyses of reaches made targets of uniform distance with the paretic and non-paretic upper limb. (**A**) Normalised wrist velocity. Shading denotes 1 SE. (**B**) Model estimates and 95%CI for Spectral Arc Length (SPARC), a measure of smoothness of the wrist velocity expressed as LI. (**C**) Trunk tilt angle was summed across from reaches made to five targets. (**D**) Model estimates and 95%CI from Mixed Models with Repeated Measures of Trunk tilt angle expressed as LI. Coloured asterisks denote significant differences from 0 from two-tailed one-sample t-tests. See Supplementary Table 6 for the number of samples per group and time and F-test statistics. *p<0.05 **p<0.01 ***p<0.001

Analysis of trunk tilt during reaching (Fig. 4C & D) indicated a main effect of Time (*F*_2,70.1_ = 3.19, *P* < 0.05), no effect of Group (*F*_1,36.9_ = 0.28, *P* = 0.6) and no Group x Time interaction (*F*_2,70_ = 0.03, *P* = 0.98). Trunk tilt LI improved between one and three months (ΔLI = -0.080, -0.014 – -0.144, *P* < 0.05), with no further improvement from three to six months post-stroke (ΔLI = 0.018, -0.0466 – 0.0824, *P* = 0.58). At all post-intervention timepoints, trunk tilt LI was less than 0 (1m LI = -0.174, -0.231 − -0.117, *P* < 0.0001, 3m LI = -0.094, -0.150 – - 0.038, *P* = 0.005, 6m LI = -0.112, -0.168 − -0.0568, *P* = 0.0001) indicating that more trunk tilt occurred when reaching with the paretic compared to the non-paretic side.

Fixed effects of baseline variables for both analyses are shown in Supplementary Table 6.

#### Stimulus-Response Curves

To quantify corticospinal excitability, TMS-derived S-R curves (Fig. 5A & B) were generated for the four muscles on both sides. For MEPsum expressed as LI (Fig. 5C), model estimates indicated there was no effect of Time (*F*_2,45.2_ = 1.50, *P* = 0.24) or Group (*F*_1,41.9_ = 0.02, *P* = 0.89), and no Group x Time interaction (*F*_2,45.3_ = 0.19, *P* = 0.83). At all post-intervention timepoints, LI was greater than 0 (1m LI = 0.54, 0.41 – 0.67, *P* < 0.0001, 3m LI = 0.48, 0.35 – 0.60, *P* < 0.0001, 6m MEP sum LI = 0.47, 0.34 – 0.59, *P* < 0.0001). These values indicate a strong bias in corticospinal excitability favouring the non-paretic side, with minimal change over time.

**Figure 5.**
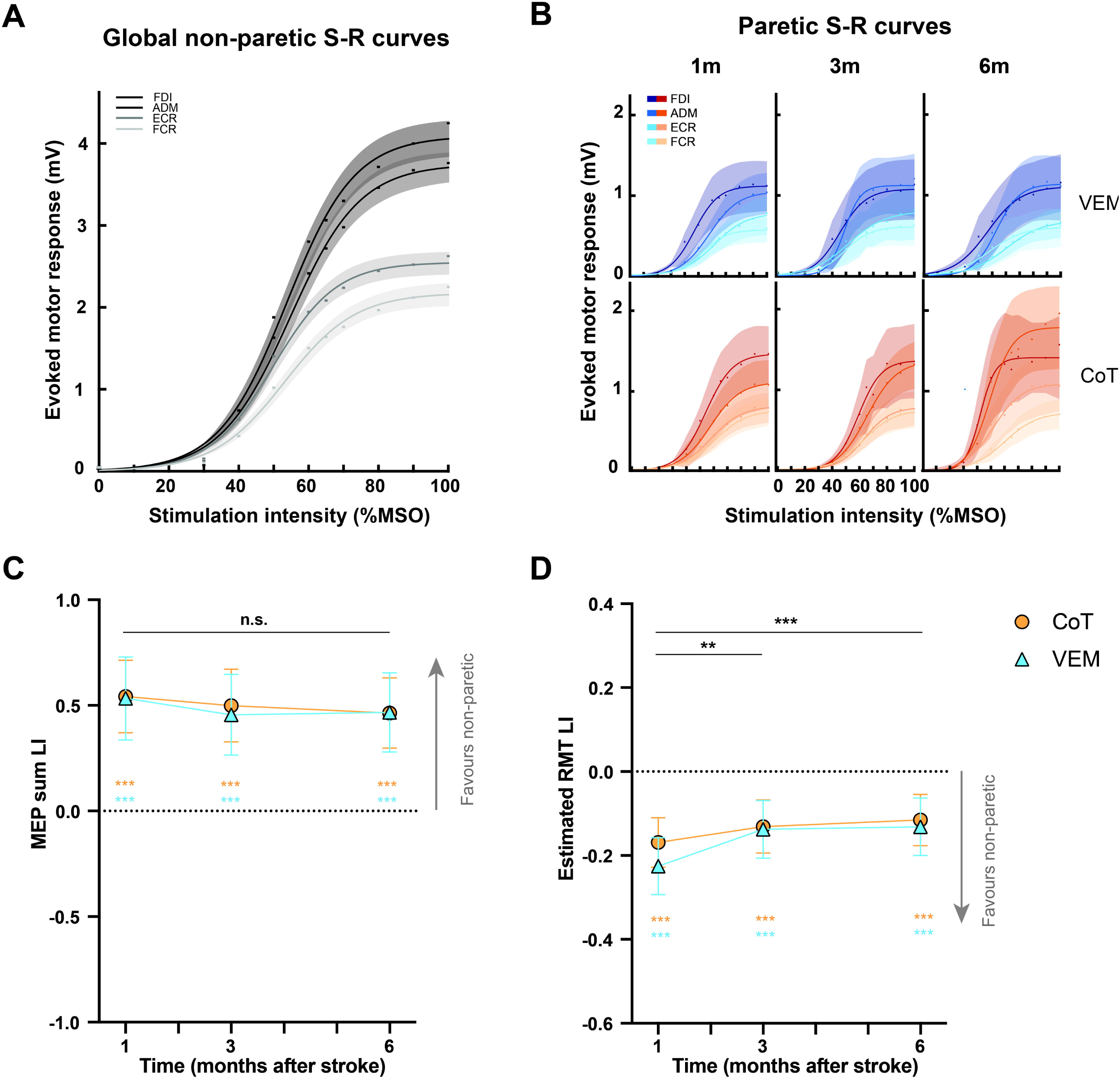
TMS-derived stimulus-response curves and measures. (**A**) Non-paretic and (**B**) paretic sides. Data points reflect group averages. Shading denotes 1 SE. Model estimates and 95%CI from Mixed Models with Repeated Measures for MEPsum which the area under S-R curves (**C**) and estimated RMT (**D**) which reflects stimulation intensity where curves first inflect, both expressed as LI. Coloured asterisks denote significant differences from 0 from two-tailed one-sample t-tests. See Supplementary Table 7 for the number of samples per group and time and F-test statistics. ADM adductor digiti minimi FDI first dorsal interosseous ECR extensor carpi radialis FCR flexor carpi radialis **p<0.01 ***p<0.001 n.s. non-significant

For eRMT derived from S-R curves expressed as LI (Fig. 5D), there was a main effect of Time (*F*_2,56.6_ = 7.60, *P* < 0.005), no effect of Group (*F*_1,39.8_ = 0.44, *P* = 0.51) and no Group x Time interaction (*F*_2,56.8_ = 0.85, *P* = 0.43). The LI increased between one and three months (ΔLI = 0.063, 0.023 – 0.10, *P* < 0.005) with no further improvement from three to six months post-stroke (ΔLI = 0.011, -0.05 – 0.03, *P* = 0.59). At all timepoints, eRMT LI was less than 0 (1m LI = -0.197, -0.24 – -0.15, *P* < 0.0001, 3m LI = -0.134, -0.18 – -0.09, *P* < 0.0001, 6m LI = -0.124, -0.17 − -0.08, *P* < 0.0001). Like MEPsum, analyses of eRMT LI indicated corticospinal pathway excitability was greater for the non-paretic side than the paretic side across all post-intervention time points.

Fixed effects of baseline variables for both analyses are shown in Supplementary Table 7.

#### Comparisons to Historical Cohort

Demographic, baseline clinical information and therapy amounts for ESPRESSo (HDHI) and Historical cohort (HC) are shown in Table 3. T-tests indicated no difference between cohorts for 3-month ARAT outcomes (HDHI = 41.2, HC = 43.3, mean difference = 2.1, -8.0 – 3.8, *P* > 0.5) or 3-month ΔFM-UE (HDHI = 20.9, HC = 21.9, mean difference = 1.0, -5.1 – 3.1, *P* > 0.6; Fig. 6A). Multiple linear regression accounting for baseline variables also indicated no differences between cohorts (Fig. 6B; see Supplement Tables S8 and S9). The HDHI cohort completed significantly more upper limb therapy than HC, who received only usual customary care (HDHI mean = 901.6 min, range 443 – 1512, HC mean = 249.3 min, range = 0 – 803, *P* < 0.0001; Fig. 6C). Pearson correlation indicated no association between ΔFM_3M_ and Total upper limb therapy time for the entire cohort (*r* = 0.085, -0.10 – 0.27, *R^2^* = 0.007, *P* = 0.3815).

**Figure 6.**
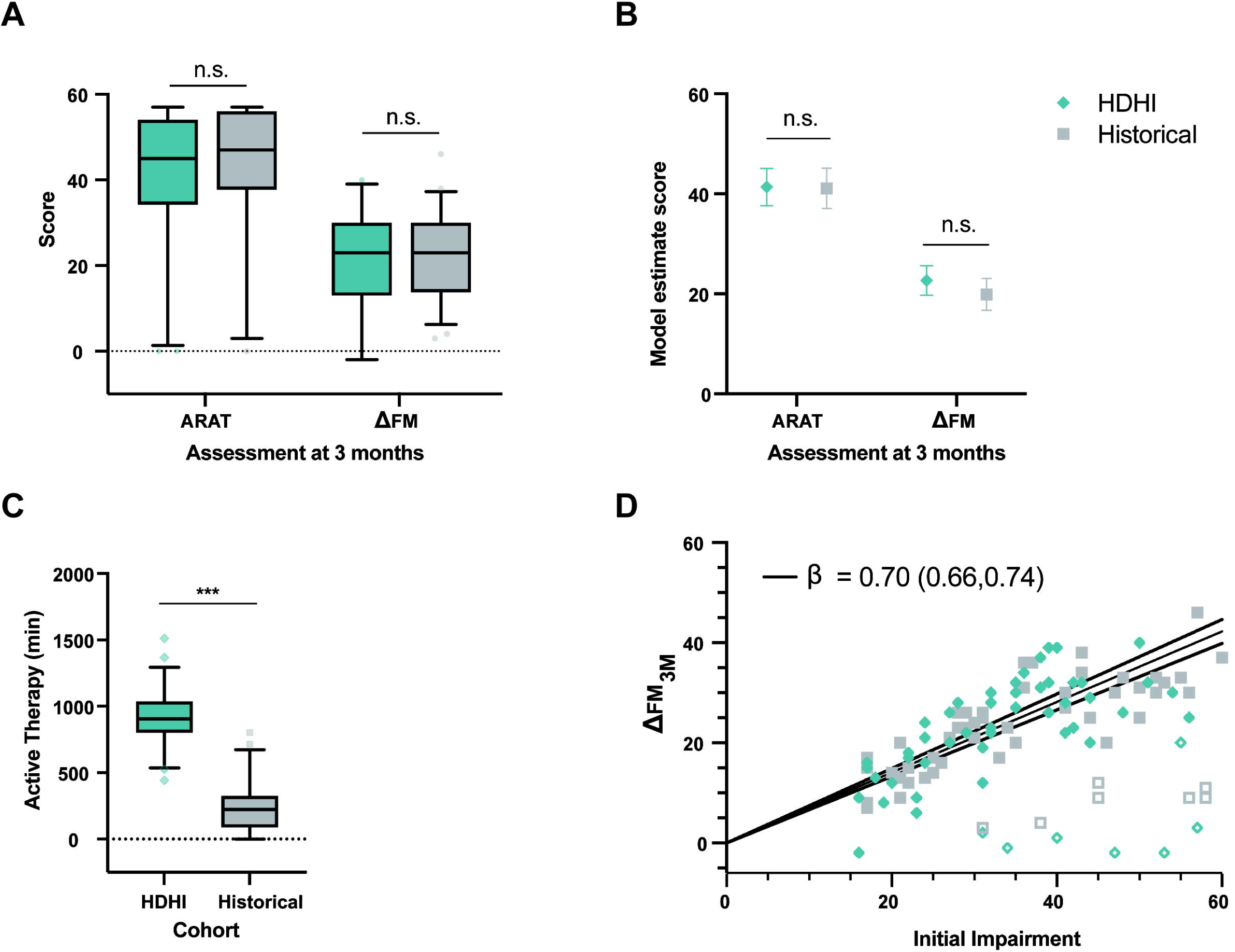
Comparing HDHI cohort (ESPRESSo *per protocol*) to Historical cohort. (**A**) Unpaired t-tests indicated no differences between HDHI (n=54) and Historical (n=54) cohorts for three-month ARAT or ΔFM. (**B**) Least squares means estimates with 95%CI from multiple linear regression, accounting for baseline variables, also indicated no difference between cohorts (see Tables S8 and S9). (**C**) An unpaired two-tailed t-test indicated active upper limb therapy time in minutes was significantly greater for HDHI than Historical cohort. (**D**) Recovery at three months (ΔFM_3M_) from linear regression with intercept forced through 0. Recovery (filled symbols) and non-recovery (unfilled symbols) phenotypes were identified based on k-means clustering, with the line of best fit (solid line), estimated slope (β) and 95%CI (dotted lines) plotted for recovery phenotype patients only (n=94, 47 per cohort). Initial impairment = 66 – FM_Baseline_. Box and whisker plots denote IQR, 5^th^ and 95^th^ percentiles. ***p<0.001

Recovery and non-recovery phenotypes were identified from k-means clustering. The approach identified two clusters (recoverers, N=94, 47 per cohort; non-recoverers, N=14, 7 per cohort; see Fig. 6D). Multiple linear regression models of ΔFM indicated a better-fitting model for recovery phenotype data compared to all patients (Supplement Tables S9 and S10). For recovery phenotype data, there was no difference in ΔFM between cohorts for (HDHI = 24.90, 22.91 – 26.89, HC = 23.40, 21.27 – 25.52, *P* = 0.280; Supplementary Table 10). The special case of proportional recovery (ΔFM_3M_ versus initial impairment) was examined with linear regression forced through the intercept (Fig. 6D). A single model fit recovery data from both cohorts better than fitting each cohort separately (*F*_1,92_ = 0.456, *P* = 0.501). Linear regression identified a line of best fit with *R^2^*= 0.64 and slope β = 0.70 (0.66 – 0.74), indicative of proportional recovery (Figure 6D).^6^

## Discussion

ESPRESSo aimed to compare the effects of two high-dose, high-intensity therapy protocols initiated within two weeks of stroke and with patients selected using biomarker guidance. There was no difference between the VEM and CoT groups at the primary endpoint, for secondary outcomes, or any neuroscientific measures. Neutral outcomes are common in early motor rehabilitation trials.^4^ The findings of the present study align with a number of recent early-phase motor rehabilitation RCTs that attempted to examine relatively high doses of upper limb therapy with no statistically significant differences between treatment and control at the primary endpoint. These include task-oriented therapy,^44^ constraint-induced movement therapy or neuromuscular stimulation,^45^ functional strength training,^46^ upper limb virtual reality training,^47^ upper limb training with Wii for 6 weeks,^48^ robot-assisted upper limb training,^49^ telerehabilitation guided conventional therapy,^50^ and a similar VEM intervention initiated later in the sub-acute phase compared to the present trial.^32^ While most of these trials did not achieve truly high-doses of therapy either, additional factors might also have contributed to the tendency for neutral findings including patient selection criteria, insufficient contrast between treatment and control arms.^4^ An earlier and smaller trial initiated later in the sub-acute phase compared VEM to CoT, both in HDHI form, and there was no difference between them. However both arms were better than a usual dose historical control group in terms of 3 month ARAT.^32^ Thus, neutral trials can occur because the novel intervention was not superior to usual care, or because the novel intervention and the control were both superior to usual care.

Despite the neutral outcome, both VEM and CoT groups made significant early hand and arm recovery as measured on FM-UE and ARAT by one-month post-stroke, with further improvement to three and six months. These gains are notable for several reasons. Motor recovery occurs rapidly within the initial few weeks after stroke, owing in large part to spontaneous biological recovery. In the present study, the largest gains in impairment and activity capacity occurred between baseline and the immediate post-intervention timepoint around one month poststroke, with more modest gains between one and six months. The fact that initial impairment is a strong predictor ΔFM and ΔARAT (Figs 6C-D; see fixed effects of baseline FM-UE in Tables S1-4) supports the contention that endogenous processes largely account for early recovery.^3,6,17^ While FM-UE and ARAT reflect levels of body function and activity respectively, the two measures tend to be highly correlated. It is also worth noting that baseline variables such as age, stroke severity (NIHSS) and the use of reperfusion therapy did not account for upper limb recovery.^29^ Also, for concordant stroke, ΔARAT at six months was ∼7 points greater than when the nondominant side was affected, a finding that is similar to a study that examined dosing of upper limb therapy at the chronic phase.^51^ This finding supports the idea that greater therapy input and motivational techniques might be needed when the nondominant side is affected. Overall, our findings underscore that upper limb recovery is strongly dictated by endogenous biological processes responsible for recovery from impairment through restitution, as well as from gains in activity capacity from compensatory mechanisms.^16^

### Kinematics, Dexterity and Corticospinal Excitability

Hemiparesis affecting the upper limb appears to be both a deficit disorder and a movement disorder, with the two related to different aspects of the interplay between a damaged CST and the reticulospinal tract (RST).^52,53^ Therefore, it is important to consider how new interventions may specifically target each deficit. In accordance with a recent consensus statement,^54^ standardised measurements of upper limb movement quality were used to assess movement smoothness, motor control and dexterity using precision grip. TMS-derived stimulus-response curves were also used to assay corticospinal excitability. Trunk tilt during reaching and hand dexterity using whole hand grasp provide assays of reticulospinal involvement. All assessments were obtained at one, three and six months post-stroke. Recovery profiles of measures assumed to rely on corticospinal function displayed early plateaus and persistent deficits compared to the non-paretic side. This observation was upheld for reaching smoothness, manual dexterity when restricted to pinch, and estimated RMT from stimulus-response curves. These measures plateaued between one and three months, at levels below (or far below) those of the non-paretic side. An early plateau was most starkly observed in MEP measures (S-R curves), which showed no change between one and six months and remained at levels far below the non-paretic side, indicating that recovery of the lower latency components of CST function in a neurophysiological sense is early, rapid and incomplete. In contrast, motor control using whole hand grasp, a functional activity that can be performed exclusively with RST, recovered to the same level as the non-paretic side by three months. These findings generally align with recent observations that lower latency components of the CST (and its associated functions) reach a plateau earlier than that made by other descending pathways, such as the RST.^23,53,55^ It is also possible that the CST itself has longer latency components that recover later and may have contributed to kinematic and dexterity outcomes.

The time course of recovery in terms of CST function was notable and surprising. There was little change in the CST input-output properties across the entire post-intervention period, as evidenced by stimulus response curves from TMS. In other words, there was no detectable change in CST gain as captured by TMS, over the same period in which recovery on ARAT and FM occurred for many participants. Curves obtained from the paretic side remaining significantly blunted relative to the non-paretic side (Fig. 5). Evidence of a functionally intact CST within a few days of stroke appears necessary, but insufficient, for achieving a favourable upper limb outcome. For example, the presence of motor-evoked potentials (MEP+ biomarker) within 5-10 days is necessary for a favourable upper limb outcome. MEP- status has very high prediction accuracy for a poor upper limb outcome.^4,8,10,28,29,56–58^ MEP+ status was confirmed before randomisation, with MEP- patients excluded, to maximise sensitivity of the trial.^28^ However, many factors can contribute to an unfavourable upper limb outcome despite MEP+ status. These may include the presence of higher-order deficits like apraxia and neglect, which result from disconnection syndromes and affect motor planning.^59^ Baseline screening ensured that all participants could engage with upper limb therapy, and the majority met protocol. Therefore, it seems unlikely that motor planning deficits can fully account for these findings. Future studies might examine whether more fine-grained analyses of MEPs can yield new insights into the poor recovery experienced by some MEP+ patients,^60^ or whether the CST may be contributing through longer latency mechanisms that are not captured by MEPs.^61^

Given the similarity in recovery and outcome of both groups, it is worth considering the factors they VEM and CoT in common. The groups were well balanced (Table 1) and had similar experiences in therapy (Table 2). Both groups received usual customary care, in terms of physical and occupational therapy, which did not differ in the amount of upper limb therapy delivered. Both groups also engaged in their respective additional therapy modalities under the direct care and supervision of experienced research therapists, with the same therapists delivering VEM and CoT. Therefore, despite the apparent contrast between VEM and CoT in terms of their emphasis on impairment versus activity capacity respectively, these common elements cannot be ignored and may have contributed to the similar outcomes. While the present trial was not powered to detect non-inferiority, it is intriguing that outcomes and recovery did not differ between VEM and CoT. Given the early and HDHI nature of both therapy modalities, these data alone do not address whether the addition of early HDHI therapy contributed to further gains in upper limb impairment and activity capacity, as have been observed at the late sub-acute phase.^32^

### Comparisons with Historical Cohort

To determine if ESPRESSo participants benefited from additional early therapy, their recovery and outcomes were compared to an historical cohort comprised of research participants receiving usual customary care only in earlier observational (non-interventional) studies undertaken previously at the same hospital. As expected, ESPRESSo participants who met protocol engaged in significantly more active upper limb therapy than those who received usual care. However, the absolute amount of therapy in terms of active therapy minutes was lower than HDHI upper limb RCTs conducted later post-stroke.^22^ This was not surprising. Patient-related factors such as weakness and fatigue clearly limited the number of minutes of active therapy achieved, as this amount increased with each week of the intervention (Table 2). This suggests that true HDHI may best be delivered later in the sub-acute phase, such that most patients are physically capable of achieving HDHI targets.

Both cohorts exhibited a remarkably stereotypical “proportional” recovery from upper limb impairment, as has been reported in many studies.^3,6–10,12,62,63^ These findings challenge the idea that simply increasing the dose or intensity of activity-based therapies at the early sub-acute phase after stroke will necessarily result in outcomes that are better than spontaneous recovery.^64^ It is worth noting that a recent Cochrane review of 21 parallel RCTs, including data from 1412 participants, compared groups who received more versus less therapy time and found that more therapy could have a positive effect on upper limb impairment compared to less therapy, but the certainty of the evidence was considered “low to very low”.^64^ Additionally, the analysis combined studies undertaken across various timepoints after stroke, blurring those that delivered therapy during versus after the critical period of SBR. The CPASS trial demonstrated benefits for ARAT outcomes when additional upper limb therapy was added to UCC at early and late sub-acute phases.^21^ However, the benefit of additional task-oriented therapy between the early sub-acute group and UCC was 4.8 points, which is below the MCID (5.7) for the ARAT. This relatively weak signal from intensive task-oriented therapy at the early sub-acute phase was one of the motivating factors for examining impairment-oriented therapy with VEM. Recovery from impairment is more closely linked to endogenous repair processes that lead to restitution, as opposed to compensation, and was expected to generalise more widely across activities.^16^ However, neither impairment nor activity capacity recovery showed a benefit of additional early therapy from levels reached in the present trial.

The findings from ESPRESSo have implications for current and future stroke rehabilitation practice. The present findings indicate that a three-fold increase in active upper limb therapy delivered during the early sub-acute phase, made little or no difference in impairment recovery or activity capacity outcome at three months. The actual time-on-task was nevertheless lower than that achieved in HDHI trials in the late sub-acute and chronic phases which achieved greater than MCID effects in the FM and ARAT. It remains an open question whether such doses, which we had aimed for when we designed the trial, would have achieved a different outcome here,^13^ but perhaps even more challenging is how such high doses would be achieved. The desire to deliver HDHI activity-based therapy early after stroke presents considerable real-world challenges that may require a “step change” in the way therapy is planned and delivered.^13,22,65^ Consistent with an earlier report, we found that VEM was feasible to deliver in an acute rehabilitation environment,^26^ provided that resource limitations, patient compliance and administrative constraints were navigated successfully. Starting within two weeks of stroke, most patients could only achieve active time on task of for about half of the extra therapy time available (i.e., 45 of 90 minutes). While patient motivation was high, fatigue increased on average by 2 points during every session, regardless of therapy modality, indicating a potential ceiling on how much and how often activity-based therapy can be delivered at the early sub-acute phase, at least across one or two sessions/day was undertaken in ESPRESSo. Even patients with high motivation and low fatigue rarely exceeded 65 of the 90 minutes available. Inactive time may reflect in part the necessary time taken to transition between game levels (VEM) or setting up different equipment when navigating between tasks (CoT). ESPRESSo illustrates the difficulty overcome patient-related factors (primarily) to deliver early HDHI therapy in the context of an RCT undertaken within a modern stroke unit in a well-resourced hospital. The likely difficulty of overcoming patient-, clinician- and system-related factors at this early time in clinical practice are worth reiterating.^27^ Future trials conducted at the early sub-acute stage might explore qualitative aspects about patient perceptions of early HDHI therapy.

Stroke rehabilitation must efficiently prioritise activities across several therapy domains, including motor (upper limb and walking), sensory, swallowing, communication and cognitive, based on the needs of individual patients. The opportunity cost for patients to engage in greater amounts of active upper limb therapy must be considered carefully by their therapy team. These different priorities might explain the large range in active upper limb therapy minutes delivered during UCC (See Table 2). The competing demands brought on by the multi-disciplinary nature of stroke rehabilitation underscore the urgent need for developing therapy modalities, including platforms like VEM, that can better accommodate a patient’s capacity and availability throughout the day.^66^ It seems likely that more upper limb therapy than currently examined could be delivered in an optimised environment. However, this does not logically lead to the conclusion that higher doses or higher intensities will necessarily interact with endogenous recovery, or that many patients will be suitable to engage with additional therapy.^13^ While the present findings did not support the hypothesis that early HDHI therapy (whether impairment-oriented or not) would enhance upper limb recovery and outcome, they do not discount the fact that meaningful gains can be made from engaging in HDHI upper limb therapy programs at the chronic phase.^67,68^ However, gains obtained at the chronic phase are inevitably smaller than the sub-acute phase, and not strictly dose-dependent.^51^

It is challenging to differentiate between gains owing to stroke rehabilitation therapy from those that are due to endogenous recovery processes. In ESPRESSo, the additional therapy did not result in improved recovery or outcome and therefore were either insufficient or unable to meaningfully interact with, or facilitate, the endogenous processes responsible for early spontaneous recovery from impairment. While the bulk of evidence from animal stroke studies indicates a benefit from early intensive activity for motor recovery,^19^ there is also some evidence for competition between endogenous and training-related processes in the very early sub-acute phase.^69^ It remains difficult to reconcile findings from animal studies with human trials undertaken in hospital settings, given that the context in which the research is conducted is so vastly different, even if the neurobiology is considered comparable.

The ESPRESSo trial has several limitations. First, the recruitment period overlapped with restrictions due to the COVID-19 pandemic, which curtailed recruitment and contributed to the study not obtaining the declared sample size. By the end of the recruitment period, about half (64 of 132) of the expected participants had been randomised to the RCT. This undoubtedly reduced the power of the study and the ability to detect between-group differences. Regardless, we randomised almost three times the number of participants in a similar trial, comparing VEM with CoT at a later time after stroke.^32^ Second, as a single-site study, patients were recruited from only one large metropolitan hospital, limiting the generalisability of the findings. Third, while imaging was undertaken at the hyper-acute phase to confirm stroke, no subsequent neuroimaging was undertaken that would have permitted a more fine-grained analysis of lesion load on descending pathways or other brain structures. Fourth, although minimisation procedures were used to balance groups for the intention-to-treat analysis, differences in baseline demographics (Table 1) may have influenced the results, perhaps owing to the smaller than anticipated sample size. Fifth, participants with more severe impairment were not always able to complete the reaching or pinch dexterity tasks, resulting in lateralisation indices that may under-estimate the actual deficit at a group level. S-R curves by contrast, were obtained at rest, and revealed larger and more persistent deficits between the paretic and non-paretic sides. Sixth, the overall amount of active upper limb therapy was lower than that delivered in trials conducted at the late sub-acute or chronic phase.^64^ The inability to achieve the maximum available minutes in terms of active therapy time may be due in part to the early start of the intervention period within two weeks of stroke, the presence of profound initial weakness, post-stroke fatigue, the relatively older age of the trial participants compared to similar trials, or a combination of these factors. However, restricting the inclusion criteria based on these factors would have reduced the generalisability of the study findings and excluded patients with potential for good upper limb recovery owing to MEP+ status.^4,8^ Finally, as mentioned, ΔARAT could not be determined for the Historical Cohort due to the absence of baseline ARAT scores.

In conclusion, the ESPRESSo RCT did not find any advantage of HDHI VEM compared to HDHI CoT therapy starting within two weeks of stroke. However, all ESPRESSo participants made considerable gains in terms of upper limb impairment and activity capacity over the sub-acute phase after stroke. By 12 weeks post-stroke, a time which typically demarcates the end of spontaneous recovery period for the upper limb, there was no measurable benefit from the additional upper limb therapy completed within the first five weeks after stroke in this study. Can early, high-dose, high-intensity therapy improve upper limb motor recovery and outcome? At present, we only know the dose that does not improve recovery outcome, and the barriers faced and overcome, in achieving that dose. While it may be tempting to deliver even greater doses of therapy to mimic the extent of motor recovery in animal studies, such an approach may not be realistic for the majority patients at the early sub-acute phase. It may simply be that HDHI interventions are best reserved until later times post-stroke. Finally, the present findings do not preclude the possibility that lower, achievable doses in the early sub-acute phase may add to higher doses at the later phase. Enhancing spontaneous recovery after stroke remains a considerable challenge.

## Data availability

The data that support the findings of this study are available from the corresponding author, upon reasonable request. The code used to analyse neuroscientific data can be found here https://github.com/wbyblow/ESPRESSo.git.

## Data Availability

https://academic.oup.com/braincomms/article-lookup/doi/10.1093/braincomms/fcag057#supplementary-data

## Acknowledgements

The authors acknowledge the contributions of the following people: Brittany Olsen-Verner, Caitlin Stewart, Catherine Hanson, Cyntia Parent, Davinia Miller, Emma Morris, Guiliana Sewell, Harry Jordan, Jess Cadenhead, Jon Bagnall, Kelly Ho, Kimberley Kar, Leanne Newall, Lindsay McGreal, Lucy Pringle, Margery Bertulfo, Marie-Claire Smith, Maya Tajitsu Jain, Olivia Norrie, Pablo Ortega-Auriol, Phoebe Ross, Samriti Sharma, Shona Mackay, Stacey Reading, Sylvain Traeger, Trish Tillson.

## Supplementary material

Supplementary material is available at *Brain Communications* online.

