## Supplementary Material for "Enhancing spontaneous recovery after stroke: A randomised controlled trial"

### Material and Methods

#### Participants

MEP status was determined by trained TMS operators as follows. Stimulus intensity was initially set to 30% maximum stimulator output (MSO) and increased in increments up to 100% MSO until MEPs were elicited. Different scalp locations around the primary motor cortex of the ipsilesional hemisphere were stimulated at each increasing stimulus intensity to try elicit MEPs. If no MEPs were elicited at 100% MSO with the participant at rest then they were stimulated at 100% MSO during a bilateral facilitation manoeuvre, which involved hugging a pillow while attempting to maximally activate their upper limb muscles bilaterally. The patient was considered MEP+ if at least two MEPs of any amplitude were observed.

#### Interventions

Qualified physiotherapists underwent in-person and video training prior to study initiation and led the delivery of the interventions. Materials, instructions and documentation were standardised between therapists, and weekly therapist meetings ensured consistency of intervention delivery. Each therapy session was delivered by a single therapist, either on the inpatient rehabilitation ward or in an outpatient clinic.

The 90 minutes of therapy time included time taken to complete the intervention, brief rests and set-up time required between activities. Therapy time did not include any substantial interruptions such as toileting or rests longer than 5 minutes. Participants with high levels of fatigue or conflicting therapy schedules were offered two blocks of 45-minute therapy sessions.

Arm weight support was provided with either the SaeboMAS (Saebo, Charlotte, North Carolina, United States) or a bespoke exoskeleton (MindMaze SA, Lausanne) to accommodate participants with severe arm paresis or fatigue. The use and choice of arm weight support was at the therapist’s discretion, based on participant comfort and fit. The level of support was adjusted for each participant at the discretion of the therapist to provide optimal movement quality (i.e., non-synergistic movement patterns within the available range of motion) while completing interventions. Support was progressively weaned as fatigue and movement quality improved.

##### Virtual Exploratory Movement Therapy (VEM)

At the time of the study, the Mindpod Dolphin games progressed up to 368 levels. The 3D movement of the paretic arm controls the creature that swims through ocean scenes, completing various goals, including chasing and eating fish, eluding attacks, and acrobatic jumping out of the water. To promote an immersive and engaging experience, a large screen display was used in an environment with dimmed lighting and high-quality audio equipment (Bose Soundbar) to project oceanic sounds and music.

##### Conventional Therapy (CoT)

Tasks were often broken down into their movement components and then practised through a gradual progression of complexity and difficulty. No more than five therapy tasks were chosen each session to promote high levels of repetition of each task. Therapists guided the level of challenge, repetitions and time taken on each task. Impairment-based intervention (e.g., scapular stability, weight-bearing, active range of motion, stretching, and strengthening) was incorporated into sessions at the discretion of therapists if it contributed towards goal attainment and task improvement. Tasks were trained using either the paretic limb or bimanually, as appropriate for successfully completing the task.

#### Blinding and Assessments

To minimise the chances of unblinding, the blinded clinical assessors worked off-site and had no contact with participants outside of scheduled follow-up assessments. Prior to each assessment, participants were instructed by one of the investigators not to speak to the assessor about the therapy they had received.

##### Reaching Kinematics

Two FLIR Blackfly S (BFS-U3-13Y3C, FLIR, Canada) cameras positioned 1.5 m apart, and angled at 60° recorded the reaching movements. The cameras were mounted on a fixed bracket system, maintaining their relative position. Movements were recorded at 170 fps using SpinView software and stored to disk.

##### TMS-derived Stimulus-Response Curves

Surface electromyography (EMG) was recorded bilaterally from the first dorsal interosseous (FDI), abductor digiti minimi (ADM), extensor carpi radialis (ECR), and flexor carpi radialis (FCR) muscles. Muscle activity was recorded using 25-mm-diameter Ag-AgCl surface electrodes (Cleartrode™ RTL, ConMed, USA) arranged using a belly-tendon montage appropriate for each muscle. Data were saved to a computer for offline analysis using Signal software (version 7.07, Cambridge Electronic Design). A common ground electrode was placed on the dorsum of the left hand. The EMG signals were amplified (x 1000), band-pass filtered (10-1000 Hz), and sampled at 2000 Hz with a CED interface system (POWER1401mkll; Cambridge Electronic Design, Cambridge, UK). EMG data were recorded for 1 s, including a 0.5 s pre-stimulus window. Rectified and smoothed pre-stimulus EMG data were visually displayed to participants, with a target line at 10 μV overlaid to assist in maintaining a resting state.

TMS intensities were randomised and delivered at an inter-stimulus interval of 6 seconds with 15% variability. Ten stimuli were delivered at the nine intensities over the marked hotspot to generate stimulus-response curves of motor evoked potential amplitudes for each muscle, as an assay of corticospinal excitability.

#### Data Processing and Dependent Measures

##### Reaching Kinematics and Smoothness

A list of 3D positions for wrist, elbow, shoulder and trunk was extracted. The reach-to-target phase of each movement was isolated based on the wrist velocity profile, which was smoothed using a Savitzky-Golay filter, using a frame size of 11 and a polynomial degree of 3. This segmentation was automated via a custom MATLAB (2023a) script and manually checked and adjusted.

The SPARC analysis was performed with a maximum cut-off frequency of 20 Hz. This frequency range was chosen to capture both normal and abnormal movement patterns. An amplitude threshold of 0.1 was used to determine the cut-off frequency, ensuring that low-amplitude fluctuations were not considered in the analysis.

##### Joint angle trajectories

The shoulder flexion profile was measured as the angle between the elbow-shoulder vector and the chest-mid hip vector. The shoulder abduction profile was measured as the angle between the elbow-shoulder vector and the chest-mid hip vector, with the elbow position projected onto the body plane defined by the chest and mid-hip positions. The elbow flexion profile was measured as the angle between the wrist-elbow vector and the shoulder-elbow vector. Finally, the trunk angle trajectory was calculated as the change in trunk angle between the initial resting position and the final position at the end of each reach, based on the chest and mid-hip vectors. To quantify the trajectory differences, the trajectories of the non-paretic hand were subtracted from those of the paretic hand, and the area under the curve of this difference was calculated.

##### Stimulus-Response Curves

EMG data were pre-processed by removing trials with excessive pre-stimulus noise. Starting at 10 μV and increasing in 5 μV increments, the minimum root mean square (RMS) value in a 50 ms pre-stimulus window that retained at least 50% of trials for all stimulus intensities for each muscle was determined. This method meant the most active trials for each muscle were removed while retaining sufficient frames for analysis. The MEP amplitude window width was 50 ms, starting 10 ms post-stimulation onset. Averages of pre-stimulus RMS and MEP amplitude for each stimulation intensity for each muscle were then calculated. The average background amplitude across all trials was used as the MEP amplitude value at 0% MSO to improve the fit of the sigmoid function. The slope of the sigmoid function was not used as an estimate of corticospinal excitability because curve-fit was inadequate for some participants, due to the small, non-modulating nature of MEPs for EMG signals on the paretic side.

#### Statistical Analyses

First-order Kenward-Roger approximation was used to estimate the denominator degrees of freedom for fixed effects. A first-order autoregressive covariance structure was used. Ordinary least squares method was used to estimate initial values for the covariance parameters. No missing data imputation was conducted. No multiple comparison adjustment was made.

### Results

Mixed models with repeated measures were used to explore effects of Group, Time and Group x Time (main article) and fixed effects of baseline variables from per protocol and intention to treat analyses (Supplementary Tables 1-7). The MMRM for ARAT and ∆ARAT included all baseline variables. For ∆FM, the models would not converge with baseline variables of Concordance and Reperfusion Therapy, and so these were removed from all subsequent analyses to prevent over-fitting. Secondary clinical outcomes of SIS and mRS for per protocol analyses are reported in the main article.

| **Supplementary Table 1. ∆ARAT Per Protocol** | | | | | | | |
| --- | --- | --- | --- | --- | --- | --- | --- |
| **MMRM parameter estimates and Type 3 Tests of Fixed Effects** | | | | | | | |
| **Effect** | **DF** | | **Estimate** | **Standard Error** | | **F value** | **p** |
| Intercept |  |  | 43.6634 | 11.9021 | |  |  |
| Age | 1 | 44.7 | -0.1463 | 0.11 | | 1.77 | 0.1905 |
| NIHSS | 1 | 44.3 | -0.635 | 0.4467 | | 2.02 | 0.1621 |
| Baseline FM-UE | 1 | 45.5 | 0.6918 | 0.2465 | | 7.88 | **0.0073** |
| Concordance (Y = 1) | 1 | 45.1 | -6.4341 | 3.342 | | 3.71 | 0.0605 |
| Reperfusion (Y = 1) | 1 | 45.2 | 3.1697 | 3.9413 | | 0.65 | 0.4255 |
| Days post stroke | 1 | 44.7 | -1.0619 | 0.6439 | | 2.72 | 0.1061 |
| Baseline ARAT | 1 | 45.8 | -1.0243 | 0.196 | | 27.31 | **<0.0001** |
| Group | 1 | 44.4 |  |  | | 0.6 | 0.442 |
| Time | 2 | 66.3 |  |  | | 31.76 | **<0.0001** |
| Group x Time | 2 | 66.2 |  |  | | 0.18 | 0.838 |
| **Least Squares Means Estimates** | | | | | | | |
|  | **Estimate** | | | **95%CI** | | | **p** |
| Time 3-1 | 5.99 | | | 3.86 | 8.12 | | **<0.0001** |
| Time 6-1 | 8.95 | | | 6.64 | 11.26 | | **<0.0001** |
| Time 6-3 | 2.96 | | | 0.74 | 5.18 | | **0.0096** |

| **Supplementary Table 2. ∆ARAT Intention to Treat** | | | | | | | |
| --- | --- | --- | --- | --- | --- | --- | --- |
| **MMRM parameter estimates and Type 3 Tests of Fixed Effects** | | | | | | | |
| **Effect** | **DF** | | **Estimate** | **Standard Error** | | **F value** | **p** |
| Intercept |  |  | 44.8219 | 11.5854 | |  |  |
| Age | 1 | 46.8 | -0.1466 | 0.1082 | | 1.84 | 0.1819 |
| NIHSS | 1 | 46.7 | -0.7064 | 0.4253 | | 2.76 | 0.1034 |
| Baseline FM-UE | 1 | 47.8 | 0.6938 | 0.2405 | | 8.31 | **0.0059** |
| Concordance (Y = 1) | 1 | 47.4 | -6.9177 | 3.2069 | | 4.65 | **0.0361** |
| Reperfusion (Y = 1) | 1 | 47.3 | 2.8533 | 3.8463 | | 0.55 | 0.4619 |
| Days post stroke | 1 | 46.8 | -1.1085 | 0.6293 | | 3.1 | 0.0847 |
| Baseline ARAT | 1 | 48.1 | -1.0192 | 0.1922 | | 28.11 | **<0.0001** |
| Group | 1 | 46.6 |  |  | | 0.49 | 0.4892 |
| Time | 2 | 68.2 |  |  | | 33.36 | **<0.0001** |
| Group x Time | 2 | 68.1 |  |  | | 0.12 | 0.8862 |
| **Least Squares Means Estimates** | | | | | | | |
|  | **Estimate** | | | **95%CI** | | | **p** |
| Time 3-1 | 6.09 | | | 3.97 | 8.2 | | **<0.0001** |
| Time 6-1 | 9.12 | | | 6.82 | 11.41 | | **<0.0001** |
| Time 6-3 | 3.03 | | | 0.82 | 5.24 | | **0.0078** |

| **Supplementary Table 3. ∆FM Per Protocol** | | | | | | | |
| --- | --- | --- | --- | --- | --- | --- | --- |
| **MMRM parameter estimates and Type 3 Tests of Fixed Effects** | | | | | | | |
| **Effect** | **DF** | | **Estimate** | **Standard Error** | | **F value** | **p** |
| Intercept |  |  | 54.1408 | 9.1061 | |  |  |
| Age | 1 | 45.8 | -0.1639 | 0.08592 | | 3.64 | 0.0627 |
| NIHSS | 1 | 45.7 | -0.4023 | 0.3563 | | 1.28 | 0.2647 |
| Baseline FM-UE | 1 | 46.4 | -0.3964 | 0.134 | | 8.76 | **0.0048** |
| Days post stroke | 1 | 46.4 | -0.9288 | 0.502 | | 3.42 | 0.0707 |
| Group | 1 | 45.3 |  |  | | 0.45 | 0.5036 |
| Time | 2 | 55.8 |  |  | | 10.81 | **0.0001** |
| Group x Time | 2 | 55.8 |  |  | | 1.18 | 0.3139 |
| **Least Squares Means Estimates** | | | | | | | |
|  | **Estimate** | | | **95%CI** | | | **p** |
| Time 3-1 | 3.09 | | | 1.35 | 4.84 | | **0.0007** |
| Time 6-1 | 4.83 | | | 2.69 | 6.96 | | **<0.0001** |
| Time 6-3 | 1.73 | | | 0.08 | 3.54 | | 0.0603 |

| **Supplementary Table 4. ∆FM Intention to Treat** | | | | | | | |
| --- | --- | --- | --- | --- | --- | --- | --- |
| **MMRM parameter estimates and Type 3 Tests of Fixed Effects** | | | | | | | |
| **Effect** | **DF** | | **Estimate** | **Standard Error** | | **F value** | **p** |
| Intercept |  |  | 54.7444 | 9.0214 | |  |  |
| Age | 1 | 47.6 | -0.1731 | 0.08509 | | 4.14 | **0.0475** |
| NIHSS | 1 | 47.7 | -0.4816 | 0.3459 | | 1.94 | 0.1702 |
| Baseline FM-UE | 1 | 48.4 | -0.3699 | 0.1316 | | 7.91 | **0.0071** |
| Days post stroke | 1 | 48.2 | -0.9537 | 0.4959 | | 3.7 | 0.0604 |
| Group | 1 | 47.3 |  |  | | 0.23 | 0.6339 |
| Time | 2 | 57.4 |  |  | | 11 | **<0.0001** |
| Group x Time | 2 | 57.4 |  |  | | 1.21 | 0.3046 |
| **Least Squares Means Estimates** | | | | | | | |
|  | **Estimate** | | | **95%CI** | | | **p** |
| Time 3-1 | 3.1 | | | 1.36 | 4.83 | | **0.0007** |
| Time 6-1 | 4.8 | | | 2.69 | 6.91 | | **<0.0001** |
| Time 6-3 | 1.7 | | | 0.1 | 3.51 | | 0.0637 |

| **Supplementary Table 5. Manual Dexterity Grasp and Pinch LI** | | | | | | | |
| --- | --- | --- | --- | --- | --- | --- | --- |
| **Grasp - MMRM parameter estimates and Type 3 Tests of Fixed Effects** | | | | | | | |
| **Effect** | **DF** | | **Estimate** | **Standard Error** | | **F value** | **p** |
| Intercept |  |  | -0.41 | 0.1004 | |  |  |
| Age | 1 | 39.8 | 0.002397 | 0.000927 | | 6.69 | **0.0135** |
| NIHSS | 1 | 42.2 | 0.01176 | 0.003912 | | 9.04 | **0.0044** |
| Baseline FM-UE | 1 | 40.5 | 0.003226 | 0.001439 | | 5.03 | **0.0305** |
| Days post stroke | 1 | 42.4 | 0.001851 | 0.005942 | | 0.1 | 0.7569 |
| Group | 1 | 41.1 |  |  | | 0.28 | 0.6014 |
| Time | 2 | 78.4 |  |  | | 3.36 | **0.0396** |
| Group x Time | 2 | 78.4 |  |  | | 0.59 | 0.5563 |
| **Least Squares Means Estimates** | | | | | | | |
|  | **Estimate** | | | **95%CI** | | | **p** |
| Time 3-1 | 0.06 | | | 0.01 | 0.11 | | **0.0305** |
| Time 6-1 | 0.06 | | | 0.01 | 0.12 | | **0.0269** |
| Time 6-3 | 0.004 | | | -0.05 | 0.06 | | 0.8836 |
| **Pinch- MMRM parameter estimates and Type 3 Tests of Fixed Effects** | | | | | | | |
| Intercept |  |  | -0.38.62 | 0.1082 | |  |  |
| Age | 1 | 38.8 | 0.00201 | 0.001003 | | 4.02 | 0.052 |
| NIHSS | 1 | 35.5 | 0.01018 | 0.004412 | | 5.33 | **0.0269** |
| Baseline FM-UE | 1 | 36.7 | 0.003161 | 0.001576 | | 4.02 | 0.0523 |
| Days post stroke | 1 | 37.7 | -0.00367 | 0.006427 | | 0.33 | 0.5712 |
| Group | 1 | 37.2 |  |  | | 0.29 | 0.5912 |
| Time | 2 | 72.9 |  |  | | 6.94 | **0.0017** |
| Group x Time | 2 | 72.8 |  |  | | 0.9 | 0.4103 |
| **Least Squares Means Estimates** | | | | | | | |
|  | **Estimate** | | | **95%CI** | | | **p** |
| Time 3-1 | 0.09 | | | 0.04 | 0.14 | | **0.0012** |
| Time 6-1 | 0.09 | | | 0.03 | 0.14 | | **0.0024** |
| Time 6-3 | -0.002 | | | -0.06 | 0.05 | | 0.9386 |

| **Supplementary Table 6. Reaching Kinematics SPARC and Trunk Tilt LI** | | | | | | | |
| --- | --- | --- | --- | --- | --- | --- | --- |
| **SPARC - MMRM parameter estimates and Type 3 Tests of Fixed Effects** | | | | | | | |
| **Effect** | **DF** | | **Estimate** | **Standard Error** | | **F value** | **p** |
| Intercept |  |  | -0.04346 | 0.04589 | |  |  |
| Age | 1 | 40.4 | -0.00062 | 0.000423 | | 2.15 | 0.1506 |
| NIHSS | 1 | 41.8 | -0.0015 | 0.001834 | | 0.67 | 0.4172 |
| Baseline FM-UE | 1 | 41.1 | 0.002153 | 0.000687 | | 9.82 | **0.0032** |
| Days post stroke | 1 | 43.2 | -0.00358 | 0.002712 | | 1.74 | 0.1936 |
| Group | 1 | 41.3 |  |  | | 1.02 | 0.319 |
| Time | 2 | 64.5 |  |  | | 6.09 | **0.0038** |
| Group x Time | 2 | 64.5 |  |  | | 0.14 | 0.8705 |
| **Least Squares Means Estimates** | | | | | | | |
|  | **Estimate** | | | **95%CI** | | | **p** |
| Time 3-1 | 0.025 | | | 0.01 | 0.04 | | **0.0017** |
| Time 6-1 | 0.02 | | | 0.01 | 0.04 | | **0.0059** |
| Time 6-3 | -0.002 | | | -0.02 | 0.01 | | 0.8006 |
| **Trunk tilt - MMRM parameter estimates and Type 3 Tests of Fixed Effects** | | | | | | | |
| Intercept |  |  | -0.09103 | 0.138 | |  |  |
| Age | 1 | 36 | -0.00078 | 0.001256 | | 0.38 | 0.54 |
| NIHSS | 1 | 36.4 | -0.00017 | 0.005476 | | 0 | 0.9747 |
| Baseline FM-UE | 1 | 38.2 | 0.002162 | 0.002055 | | 1.11 | 0.2992 |
| Days post stroke | 1 | 38.3 | -0.00427 | 0.008156 | | 0.27 | 0.6035 |
| Group | 1 | 36.9 |  |  | | 0.28 | 0.5993 |
| Time | 2 | 70.1 |  |  | | 3.19 | **0.0474** |
| Group x Time | 2 | 70 |  |  | | 0.03 | 0.9739 |
| **Least Squares Means Estimates** | | | | | | | |
|  | **Estimate** | **95%CI** | | | | **p** | |
| Time 3-1 | 0.080 | 0.14 | | | | 0.01 | **0.0171** |
| Time 6-1 | 0.062 | 0.13 | | | | -0.01 | 0.0713 |
| Time 6-3 | -0.018 | 0.05 | | | | -0.08 | 0.5808 |

| **Supplementary Table 7. Neurophysiology MEPsum and eRMT LI** | | | | | | | |
| --- | --- | --- | --- | --- | --- | --- | --- |
| **MEPsum - MMRM parameter estimates and Type 3 Tests of Fixed Effects** | | | | | | | |
| **Effect** | **DF** | | **Estimate** | **Standard Error** | | **F value** | **p** |
| Intercept |  |  | 0.3898 | 0.381 | |  |  |
| Age | 1 | 40.1 | 0.001938 | 0.003523 | | 0.3 | 0.5853 |
| NIHSS | 1 | 42 | -0.00803 | 0.01459 | | 0.3 | 0.5851 |
| Baseline FM-UE | 1 | 41.1 | -0.00521 | 0.005573 | | 0.87 | 0.3557 |
| Days post stroke | 1 | 42.3 | 0.02829 | 0.02234 | | 1.6 | 0.2124 |
| Group | 1 | 41.5 |  |  | | 0.02 | 0.8923 |
| Time | 2 | 45.2 |  |  | | 1.5 | 0.235 |
| Group x Time | 2 | 45.3 |  |  | | 0.16 | 0.8283 |
| **Estimated RMT- MMRM parameter estimates and Type 3 Tests of Fixed Effects** | | | | | | | |
| Intercept |  |  | -0.1043 | 0.1286 | |  |  |
| Age | 1 | 37.9 | -0.00079 | 0.001187 | | 0.44 | 0.5089 |
| NIHSS | 1 | 40.5 | 0.01201 | 0.004923 | | 5.95 | **0.0192** |
| Baseline FM-UE | 1 | 39.2 | 0.001662 | 0.001876 | | 0.78 | 0.3812 |
| Days post stroke | 1 | 41.2 | -0.01754 | 0.007557 | | 5.38 | **0.0253** |
| Group | 1 | 39.8 |  |  | | 0.44 | 0.5126 |
| Time | 2 | 56.6 |  |  | | 7.6 | **0.0012** |
| Group x Time | 2 | 56.8 |  |  | | 0.85 | 0.4307 |
| **Least Squares Means Estimates** | | | | | | | |
|  | **Estimate** | | | **95%CI** | | | **p** |
| Time 3-1 | 0.06 | | | 0.02 | 0.1 | | **0.0025** |
| Time 6-1 | 0.07 | | | 0.03 | 0.12 | | **0.0008** |
| Time 6-3 | 0.01 | | | 0.03 | 0.05 | | 0.5894 |

#### During Intervention – Cognitive Efficiency

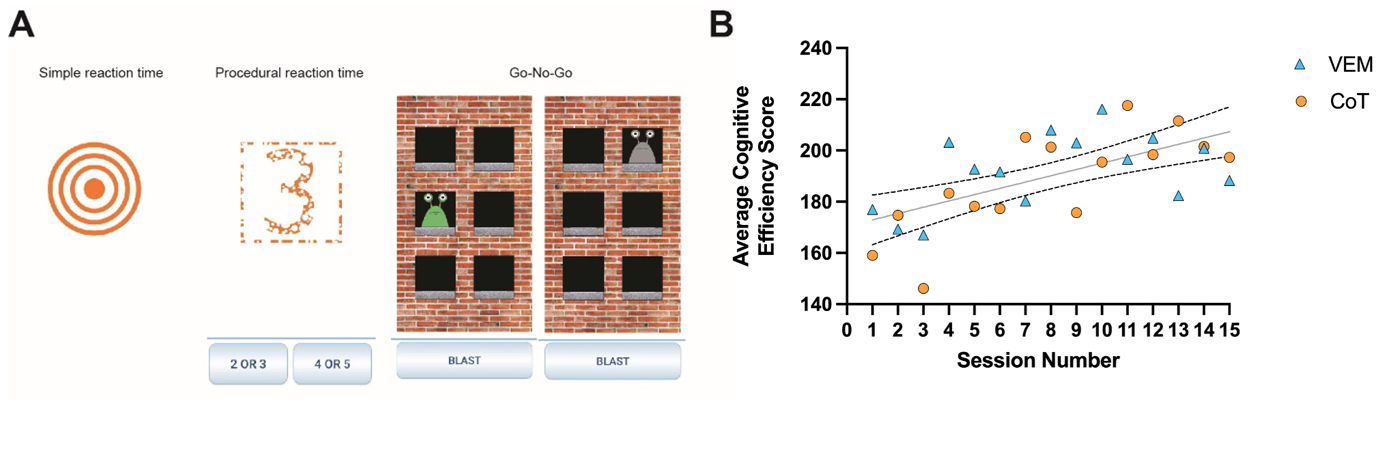

**Figure S1** Cognitive efficiency was determined from performance of three reaction time (RT) tasks (A) completed on a smartphone running the DANA Brain Vital app during rest breaks within each of the 15 therapy sessions, which comprised the interventions. (B) Linear regression indicated participants across both VEM (N=24) and CoT (N=30) from per protocol analysis tended toward greater cognitive efficiency (faster RT) over therapy sessions. Each data point reflects the mean score of the group. Solid Line y = 2.46x + 170.5, Dashed lines = 95%CI, *F* _2,26_ = 1.61, *P* = 0.28, indicates one curve for both groups.

#### During Intervention – Enjoyment (VEM Group)

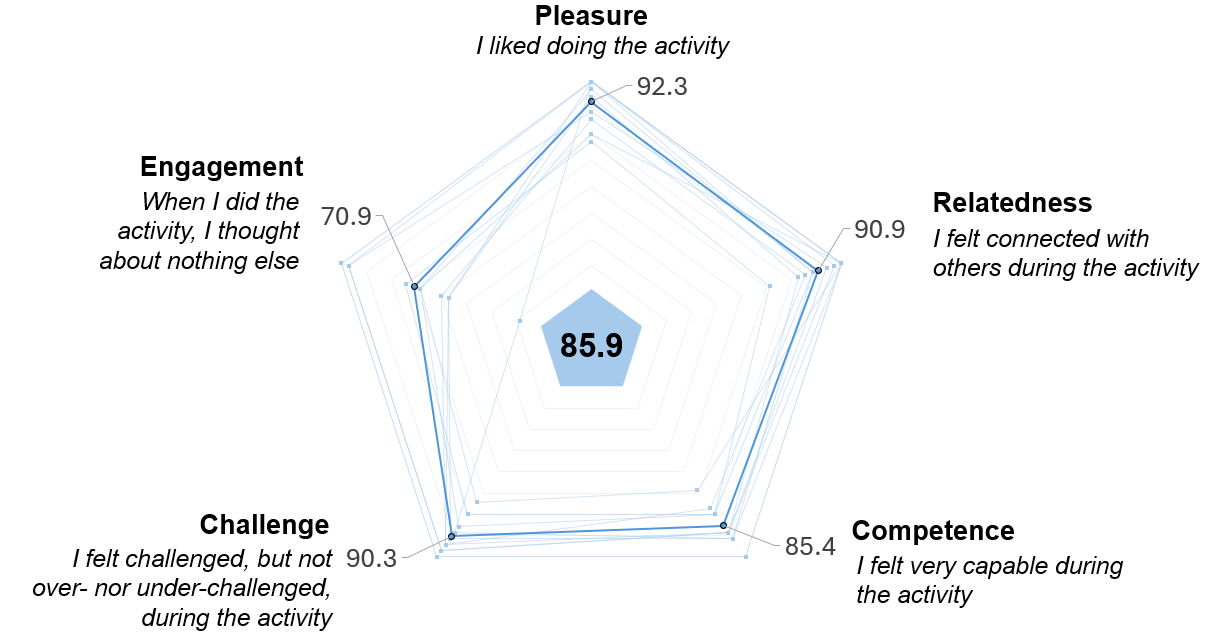

**Figure S2** A subset of participants (N=10) rated the enjoyment of the Mindpod Dolphin Platform used to deliver VEM. Each line reflects individual participant data on 5 dimensions of the ENJOY scale, along with group mean and grand average score (maximum score = 100).

#### Comparison to Historical Cohort (HC)

Unpaired t-tests and multiple linear regression models indicated no differences at three months between HDHI versus Historical cohorts. Multiple regression model estimates with baseline variables are shown in Supplementary Tables 8-10.

| **Supplementary Table 8. Multiple linear regression of 3m ARAT for HDHI and Historical (all patients)** | | | | | |
| --- | --- | --- | --- | --- | --- |
| **ARAT at 3 months** |  | **DF** | | **F** | **p** |
| **Model** |  | 7 | 107 | 11.96 | **<0.0001** |
|  | **R^2^** | **CV** | | **RMSE** | **Mean** |
|  | 0.46 | 27.83 | | 11.76 | 42.26 |
| **Parameter** | **Estimate** | **Std Err** | | **t value** | **p** |
| Intercept | 51.83 | 7.80 | | 6.65 | **<0.0001** |
| Age | -0.19 | 0.08 | | -2.51 | **0.0137** |
| NIHSS | -1.01 | 0.32 | | -3.11 | **0.0024** |
| Baseline FM-UE | 0.49 | 0.11 | | 4.5 | **<0.0001** |
| Concordance (Y = 1) | -2.86 | 2.38 | | 1.2 | 0.2332 |
| Reperfusion (Y = 1) | 2.69 | 3.00 | | 0.9 | 0.3711 |
| Days Post Stroke | -1.11 | 0.51 | | -2.19 | **0.0312** |
| Cohort (HC = 1) | 0.27 | 2.62 | | 0.1 | 0.9184 |

| **Supplementary Table 9. Multiple linear regression model of 3m ∆FM for HDHI and Historical (all patients)** | | | | | |
| --- | --- | --- | --- | --- | --- |
| **∆FM at 3 months** |  | **DF** | | **F** | **p** |
| **Model** |  | 7 | 107 | 5.83 | **<0.0001** |
|  | **R^2^** | **CV** | | **RMSE** | **Mean** |
|  | 0.29 | 43.55 | | 9.34 | 21.44 |
| **Parameter** | **Estimate** | **Std Err** | | **t Value** | **p** |
| Intercept | 52.93 | 6.19 | | 8.55 | **<0.0001** |
| Age | -0.13 | 0.06 | | -2.2 | **0.0302** |
| NIHSS | -0.39 | 0.26 | | -1.53 | 0.1301 |
| Baseline FM-UE | -0.45 | 0.09 | | -5.24 | **<.0001** |
| Concordance (Y = 1) | -2.26 | 1.89 | | 1.2 | 0.2344 |
| Reperfusion (Y = 1) | -0.11 | 2.38 | | 0.05 | 0.9641 |
| Days Post Stroke | -1.22 | 0.40 | | -3.03 | **0.0031** |
| Cohort (HC = 1) | 2.79 | 2.08 | | 1.34 | 0.184 |

| **Supplementary Table 10. Multiple linear regression of 3m ∆FM for recovery phenotype** | | | | | |
| --- | --- | --- | --- | --- | --- |
| **∆FM at 3 months** |  | **DF** | | **F** | **p** |
| **Model** |  | 7 | 93 | 20.71 | **<0.0001** |
|  | **R^2^** | **CV** | | **RMSE** | **Mean** |
|  | 0.63 | 24.27 | | 5.78 | 23.81 |
| **Parameter** | **Estimate** | **Std Err** | | **t Value** | **p** |
| Intercept | 51.38 | 3.93 | | 13.07 | **<0.0001** |
| Age | -0.07 | 0.04 | | -1.63 | 0.1073 |
| NIHSS | -0.04 | 0.17 | | -0.25 | 0.805 |
| Baseline FM-UE | -0.63 | 0.06 | | -11.16 | **<0.0001** |
| Concordance (Y = 1) | -0.34 | 1.25 | | -0.27 | 0.7849 |
| Reperfusion (Y = 1) | -1.26 | 1.60 | | -0.78 | 0.4351 |
| Days Post Stroke | -0.47 | 0.28 | | -1.67 | 0.0979 |
| Cohort (HC = 1) | 1.50 | 1.39 | | 1.09 | 0.2803 |
